# SARS-CoV-2 shedding dynamics across the respiratory tract, sex and disease severity for adult and pediatric COVID-19

**DOI:** 10.1101/2021.02.17.21251926

**Authors:** Paul Z. Chen, Niklas Bobrovitz, Zahra Premji, Marion Koopmans, David N. Fisman, Frank X. Gu

## Abstract

**Background:** SARS-CoV-2 shedding dynamics influence the risk of transmission and clinical manifestations of COVID-19. Yet, the relationships between SARS-CoV-2 shedding dynamics in the upper (URT) and lower respiratory tract (LRT) and age, sex and COVID-19 severity remain unclear.

**Methods:** Using systematic review, we developed a dataset of case characteristics (age, sex and COVID-19 severity) and quantitative respiratory viral loads (rVLs). We then conducted stratified analyses to assess SARS-CoV-2 shedding across disease course, COVID-19 severity, the respiratory tract, sex and age groups (aged 0 to 17 years, 18 to 59 years, and 60 years or older).

**Results:** The systematic dataset included 1,266 adults and 136 children with COVID-19. In the URT, adults with severe COVID-19 had higher rVLs at 1 day from symptom onset (DFSO) than adults (*P* = 0.005) or children (*P* = 0.017) with nonsevere illness. Between 1-10 DFSO, severe adults had comparable rates of SARS-CoV-2 clearance from the URT as nonsevere adults (*P* = 0.479) and nonsevere children (*P* = 0.863). In the LRT, severe adults showed higher rVLs post-symptom onset than nonsevere adults (*P* = 0.006). In the analyzed period (4-10 DFSO), severely affected adults had no significant trend in SARS-CoV-2 clearance from LRT (*P* = 0.105), whereas nonsevere adults showed a clear trend (*P* < 0.001). After stratifying for disease severity, sex and age (including child vs. adult) were not predictive of the duration of respiratory shedding. The estimated accuracy for using URT shedding as a prognostic indicator for COVID-19 severity was up to 65%, whereas it was up to 81% for LRT shedding.

**Conclusions:** High, persistent LRT shedding of SARS-CoV-2 characterized severe COVID-19 in adults. After symptom onset, severe cases tended to have slightly higher URT shedding than their nonsevere counterparts. Disease severity, rather than age or sex, predicted SARS-CoV-2 kinetics. LRT specimens more accurately prognosticate COVID-19 severity than do URT specimens.

**Funding:** Natural Sciences and Engineering Research Council of Canada (NSERC) Discovery Grant, NSERC Senior Industrial Research Chair and the Toronto COVID-19 Action Fund.

## INTRODUCTION

As of 13 May 2021, the coronavirus disease 2019 (COVID-19) pandemic has caused more than 160.5 million infections and 3.3 million deaths globally (Dong, Du, & Gardner, 2020). The clinical spectrum of COVID-19, caused by severe acute respiratory syndrome coronavirus 2 (SARS-CoV-2), is wide, ranging from asymptomatic infection to fatal disease. Risk factors for severe illness and death include age, sex, smoking and comorbidities, such as obesity, hypertension, diabetes and cardiovascular disease (Onder, Rezza, & Brusaferro, 2020; Tartof et al., 2020; Zhou et al., 2020). Cases that deteriorate into severe disease do so, on median, 10 days from symptom onset (DFSO) (Berlin, Gulick, & Martinez, 2020; Zhou et al., 2020). Emerging evidence indicates that age and sex differences in innate, cross-reactive and adaptive immunity facilitate the higher risks observed in older and male cases (Ng et al., 2020; Pierce et al., 2020; Rydyznski et al., 2020; Takahashi et al., 2020). Robust immune responses putatively mediate nonsevere illness, in part, by controlling the replication of SARS-CoV-2 (Lucas et al., 2021; Lucas et al., 2020).

As a respiratory virus, the shedding dynamics of SARS-CoV-2 in the upper (URT) and lower respiratory tract (LRT) provide insight into clinical and epidemiological factors. URT viral load has been associated with transmission risk, duration of infectiousness, disease severity and mortality (Fu et al., 2021; Magleby et al., 2020; Marks et al., 2021; Pujadas et al., 2020; van Kampen et al., 2021; Westblade et al., 2020; Wolfel et al., 2020). Key questions, however, remain. While chest computed tomography (CT) evidence of viral pneumonitis suggests pulmonary replication in most symptomatic cases (Bernheim et al., 2020), the LRT kinetics of SARS-CoV-2, especially as related to disease severity, remain unknown. The relationships between age, sex and disease severity on shedding dynamics are unclear, particularly for children. Moreover, the accuracy of viral load in predicting COVID-19 severity is poorly understood, with conflicting results for shedding dynamics based on analyses of low sample numbers (Argyropoulos et al., 2020; Lucas et al., 2020; Pujadas et al., 2020; Silva et al., 2021; Walsh et al., 2020; Westblade et al., 2020).

For insight into these questions, we conducted a systematic review on SARS-CoV-2 quantitation from respiratory specimens and developed a large, diverse dataset of respiratory viral loads (rVLs) and individual case characteristics. Stratified analyses then assessed SARS-CoV-2 shedding dynamics across the respiratory tract, age, sex and COVID-19 severity.

## RESULTS

### Overview of contributing studies

The systematic search (***Figure 1—Source Data 1 to 5***) identified 5,802 deduplicated results. After screening and full-text review, 26 studies met the inclusion criteria (Methods), and data were collected for individually reported specimens of known type with known DFSO and for COVID-19 cases with known age, sex or severity (***Figure 1***). From 1,402 COVID-19 cases, we collected 1,915 quantitative specimen measurements (viral RNA concentration in a respiratory specimen) of SARS-CoV-2 (***Table 1***) and used them to estimate respiratory viral loads (rVLs, viral RNA concentration in the respiratory tract) (Methods, ***Figure 1—Figure Supplement 1***). For pediatric cases, the search found only nonsevere infections and URT specimen measurements. ***Appendix Tables 1 and 2*** summarize the characteristics of contributing studies, of which 18 had low risk of bias according to the modified JBI critical appraisal checklist.

**Figure 1.**
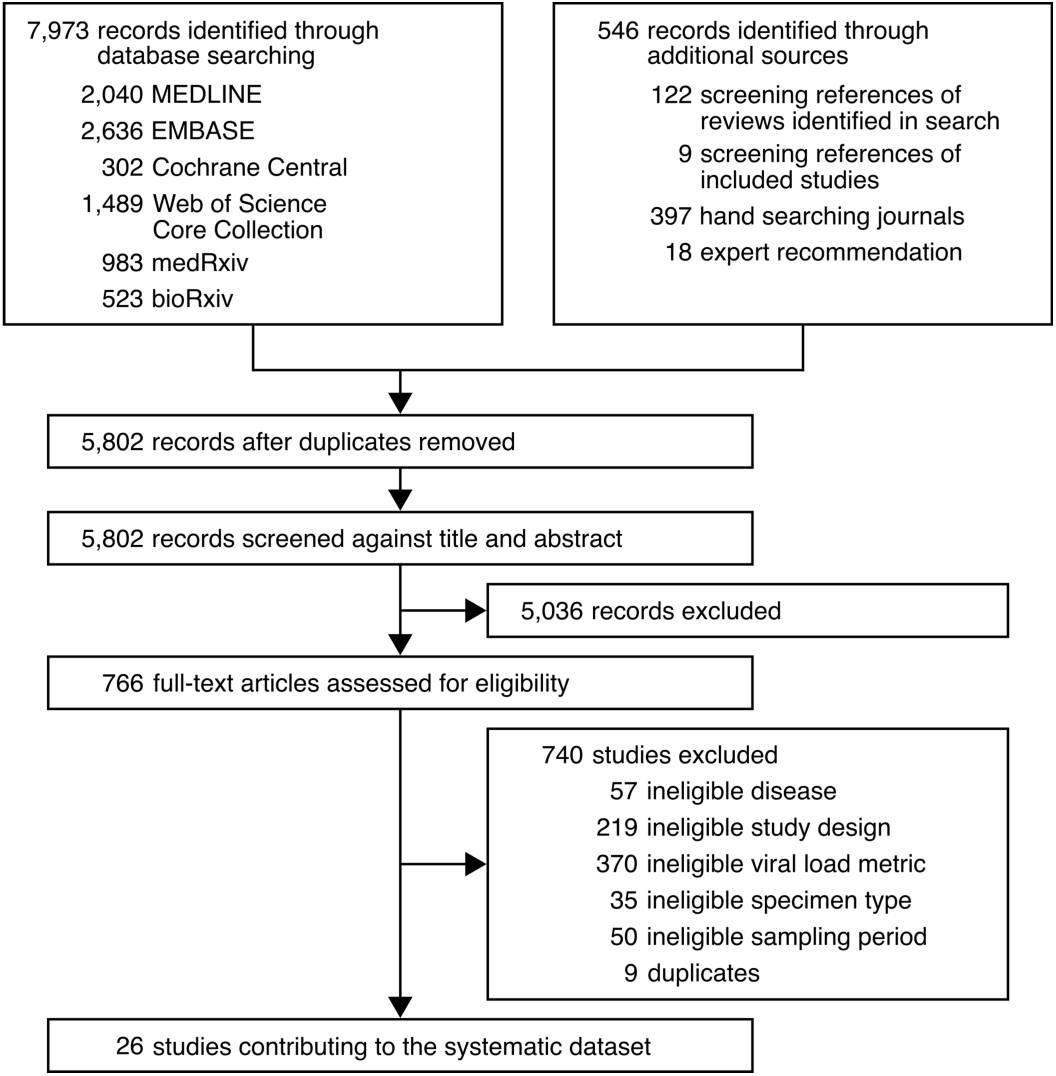
Study selection.

**Table 1.** Characteristics of adult and pediatric COVID-19 cases in the systematic dataset

|  | **Adult** | **Pediatric** |
| --- | --- | --- |
| Cases, *n* | 1 266 | 136 |
| URT specimens, *n* | 1 513 | 192 |
| LRT specimens, *n* | 210 | 0 |
| Mean age (SD), *y* | 51.8 (18.0) | 8.7 (5.3) |
| Male, *n* (%) | 528 (44.0) | 63 (52.5) |
| Disease severity, *n* (%) |  |  |
| Asymptomatic | 2 (0.2) | 5 (3.7) |
| Mild | 710 (57.5) | 112 (83.6) |
| Moderate | 178 (14.4) | 17 (12.7) |
| Severe | 167 (13.5) | 0 (0.0) |
| Critical | 178 (14.4) | 0 (0.0) |

Adult cases were those aged 18 y or older, while pediatric cases were those aged younger than 18
y. Upper respiratory tract = URT. Lower respiratory tract = LRT.

### URT shedding of SARS-CoV-2 for adult COVID-19

To interpret the complex interplay between SARS-CoV-2 shedding dynamics and age, sex and COVID-19 severity, we stratified our systematic dataset into age, sex and severity groups and then conducted a series of regression analyses. For adult COVID-19, regression analysis showed that the mean URT rVL at 1 DFSO was significantly greater (*P* for intercept = 0.005) for severely infected cases (8.28 [95% CI: 7.71-8.84] log_10_ copies/ml) than nonsevere ones (7.45 [95% CI: 7.26-7.65] log_10_ copies/ml) (***Figure 2, A and D***). Meanwhile, these groups showed comparable URT dynamics post-symptom onset (*P* for interaction = 0.479), as severe adult cases tended to cleared SARS-CoV-2 from the URT at -0.31 (95% CI: -0.40 to -0.22) log_10_ copies/ml day^-1^ while nonsevere ones did so at -0.28 (95% CI: -0.32 to -0.24) copies/ml day^-1^ (***Figure 2, A and E***). For severe cases, the estimated mean duration of URT shedding (down to 0 log_10_ copies/ml) was 27.5 (95% CI: 21.2-33.8) DFSO; it was 27.9 (95% CI: 24.4-31.3) DFSO for nonsevere cases.

**Figure 2.**
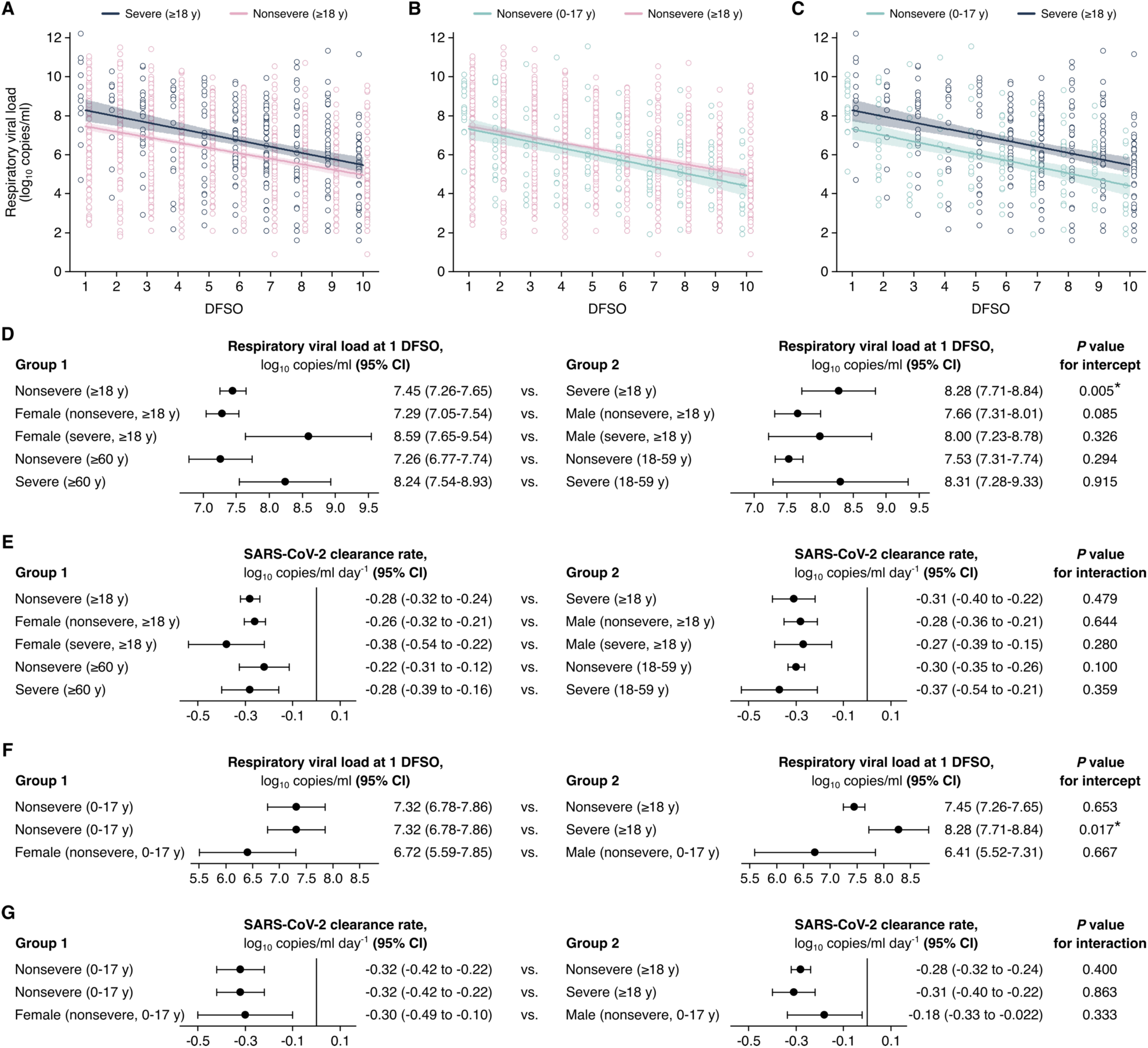
**Comparison of SARS-CoV-2 shedding in the URT across disease severity, sex and age groups**. (**A** to **C**). Upper respiratory tract (URT) shedding for severe and nonsevere adult (aged 18 y or older) COVID-19 (A), for nonsevere pediatric (aged 0-17 y) and nonsevere adult COVID-19 (B) and for nonsevere pediatric and severe adult COVID-19 (C). Open circles represent rVL data and were offset from their DFSO for visualization. Lines and bands show regressions and their 95% CIs, respectively. (**D** and **E**) Comparisons of URT shedding levels at 1 day from symptom onset (DFSO) (D) and URT shedding dynamics (E) between severity, age and sex groups for COVID-19. (**F** and **G**) Comparisons of URT shedding levels at 1 DFSO (F) and URT shedding dynamics (G) between pediatric and adult groups for COVID-19. The black line in (E) and (G) depicts 0, the threshold for no significant trend in SARS-CoV-2 clearance. Regression analyses determined *P* values and compared shedding levels and dynamics between the two groups in each row.

After stratifying adults for disease severity, our analyses showed no significant differences in URT shedding levels or dynamics between sex or age groups (***Figure 2, D and E, Figure 2— Figure Supplement 1***). For severe disease, male and female cases had comparable mean rVLs at 1 DFSO (*P* for intercept = 0.326) and rate of viral clearance (*P* for interaction = 0.280). Similarly, for nonsevere illness, male and female cases had no significant difference in mean rVL at 1 DFSO (*P* for intercept = 0.085) or URT dynamics (*P* for interaction = 0.644). For nonsevere illness, younger and older adults had no significant difference in URT shedding levels at 1 DFSO (*P* for intercept = 0.294) or post-symptom-onset dynamics (*P* for interaction = 0.100). For severe disease, the adult age groups showed similar mean rVLs at 1 DFSO (*P* for intercept = 0.915) and rates of viral clearance (*P* for interaction = 0.359).

### URT shedding of SARS-CoV-2 for pediatric COVID-19

For pediatric COVID-19, regression estimated, in the URT, the mean rVL at 1 DFSO to be 7.32 (95% CI: 6.78-7.86) log_10_ copies/ml and SARS-CoV-2 clearance rate as -0.32 (95% CI: -0.42 to -0.22) log_10_ copies/ml day^-1^ (***Figure 2, F and G***). Both estimates were comparable between the sexes for children (***Figure 2—Figure Supplement 1***). The estimated mean duration of URT shedding (down to 0 log_10_ copies/ml) was 22.6 (95% CI: 17.0-28.1) DFSO for children with COVID-19.

Between pediatric cases, who had nonsevere illness in our dataset, and adults with nonsevere illness, both URT shedding at 1 DFSO (*P* for intercept = 0.653) and URT dynamics (*P* for interaction = 0.400) were similar (***Figure 2, B, F and G***). Conversely, URT shedding at 1 DFSO was greater for severely affected adults when compared to children with nonsevere disease (*P* for intercept = 0.017), but URT dynamics remained similar (*P* for interaction = 0.863) (***Figure 2, C, F and G***).

### LRT shedding of SARS-CoV-2 for adult COVID-19

For adults, our analyses showed that high, persistent LRT shedding of SARS-CoV-2 was associated with severe COVID-19 but not nonsevere illness (***Figure 3A***). At the initial day in our analyzed period (4 DFSO), the mean rVL in the LRT of severe cases (8.42 [95% CI: 7.67-9.17] log_10_ copies/ml) was significantly greater (*P* for intercept = 0.006) than that of nonsevere cases (6.82 [95% CI: 5.95-7.69] log_10_ copies/ml) (***Figure 3D***). Between severities, the difference in LRT clearance rates was marginally above the threshold for statistical significance (*P* for interaction = 0.053). Nonetheless, severe cases had persistent LRT shedding, with no significant trend in SARS-CoV-2 clearance up to 10 DFSO (-0.14 [95% CI: -0.32 to 0.030] log_10_ copies/ml day^-1^, *P* = 0.105), whereas nonsevere cases rapidly cleared the virus from the LRT (-0.41 [95% CI: -0.64 to -0.19] log_10_ copies/ml day^-1^, *P* < 0.001) (***Figure 3E***). For nonsevere cases, the estimated mean duration of LRT shedding (down to 0 log_10_ copies/ml) was 20.4 (95% CI: 13.2-27.7) DFSO.

**Figure 3.**
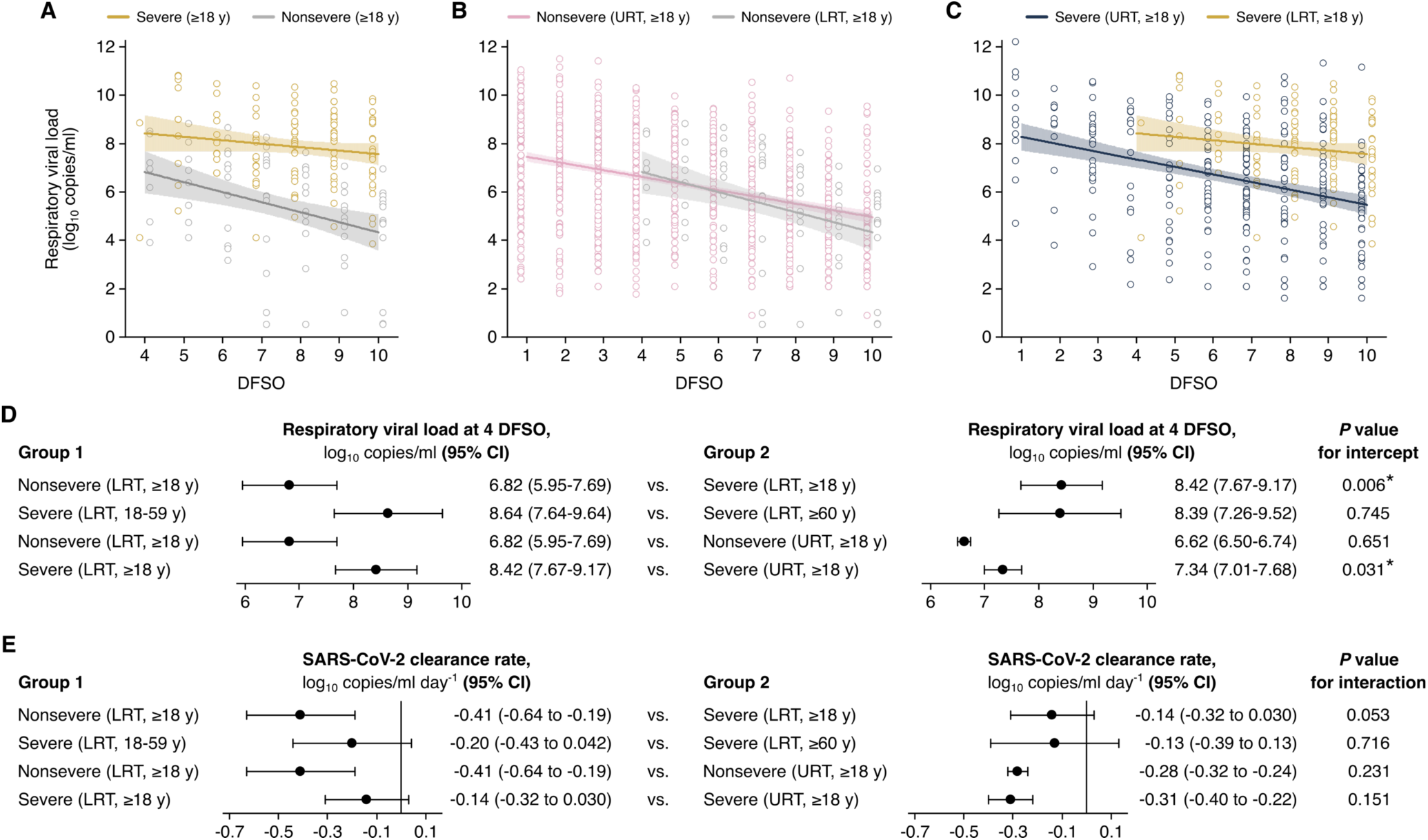
**Comparison of SARS-CoV-2 shedding in the LRT across disease severity and age groups and the respiratory tract**. (**A** to **C**). Shedding in the lower respiratory tract (LRT) for severe and nonsevere adult (aged 18 y or older) COVID-19 (A), in the LRT and upper respiratory tract (URT) nonsevere adult COVID-19 (B) and in the LRT and URT severe adult COVID-19 (C). Open circles represent rVL data and were offset from their DFSO for visualization. Lines and bands show regressions and their 95% CIs, respectively. (**D** and **E**) Comparisons of shedding levels at 4 day from symptom onset (DFSO) (D) and URT shedding dynamics (E) between severity and age groups in the LRT and between the LRT and URT. The black line in (E) depicts 0, the threshold for no significant trend in SARS-CoV-2 clearance. Regression analyses determined *P* values and compared shedding levels and dynamics between the two groups in each row.

For severe COVID-19, regression analysis showed, in the LRT, comparable mean rVLs at 4 DFSO between younger and older adults (*P* for intercept = 0.745) (***Figure 3D***). For severe cases, both age groups also showed persistent LRT shedding in the analyzed period: younger adults (- 0.20 [95% CI: -0.32 to 0.042] log_10_ copies/ml day^-1^, *P* = 0.105) and older adults (-0.13 [95% CI: -0.39 to 0.13] log_10_ copies/ml day^-1^, *P* = 0.316) both had no significant trend in SARS-CoV-2 clearance (***Figure 3E, Figure 3—Figure Supplement 1A***). Likewise, severely affected male cases had no significant trend in LRT shedding (0.001 [95% CI: -0.16 to 0.19] log_10_ copies/ml day^-1^, *P* = 0.988) (***Figure 3—Figure Supplement 1B***). The female group included few samples, and statistically analyses were not conducted (**Appendix Table 3**).

Interestingly, nonsevere cases showed similar SARS-CoV-2 shedding between the URT and LRT (***Figure 3B***), whereas severe cases shed greater and longer in the LRT than the URT (***Figure 3C***). At 4 DFSO, the URT rVL of nonsevere adults was 6.62 (95% CI: 6.50-6.74) log_10_ copies/ml, which was not different from the LRT rVL of nonsevere adults (*P* for intercept = 0.651). In contrast, for severe adults, the rVL at 4 DFSO was significantly lower in the URT (7.34 [95% CI: 7.01-7.68] log_10_ copies/ml) than the LRT (*P* for intercept = 0.031) (***Figure 3D***).

### Heterogeneity in URT shedding of SARS-CoV-2

While regression analyses compared mean shedding levels and dynamics, we fitted rVLs to Weibull distributions to assess the heterogeneity in rVL. Both severe and nonsevere adult COVID-19 showed comparably broad distributions of URT shedding throughout disease course (***Figure 4A***). For severe disease, the standard deviation (SD) of rVL was 1.86, 2.34, 1.89 and 1.90 log_10_ copies/ml at 2, 4, 7 and 10 DFSO, respectively. For nonsevere illness, these SDs were 2.08, 1.90, 1.89 and 1.96 log_10_ copies/ml, respectively. Notably, our distribution analyses indicated that the top 2-9% of adults with COVID-19 harbored 80% of the SARS-CoV-2 copies in the URT on each DFSO (***Figure 4—Figure Supplement 1, A to D***).

**Fig. 4.**
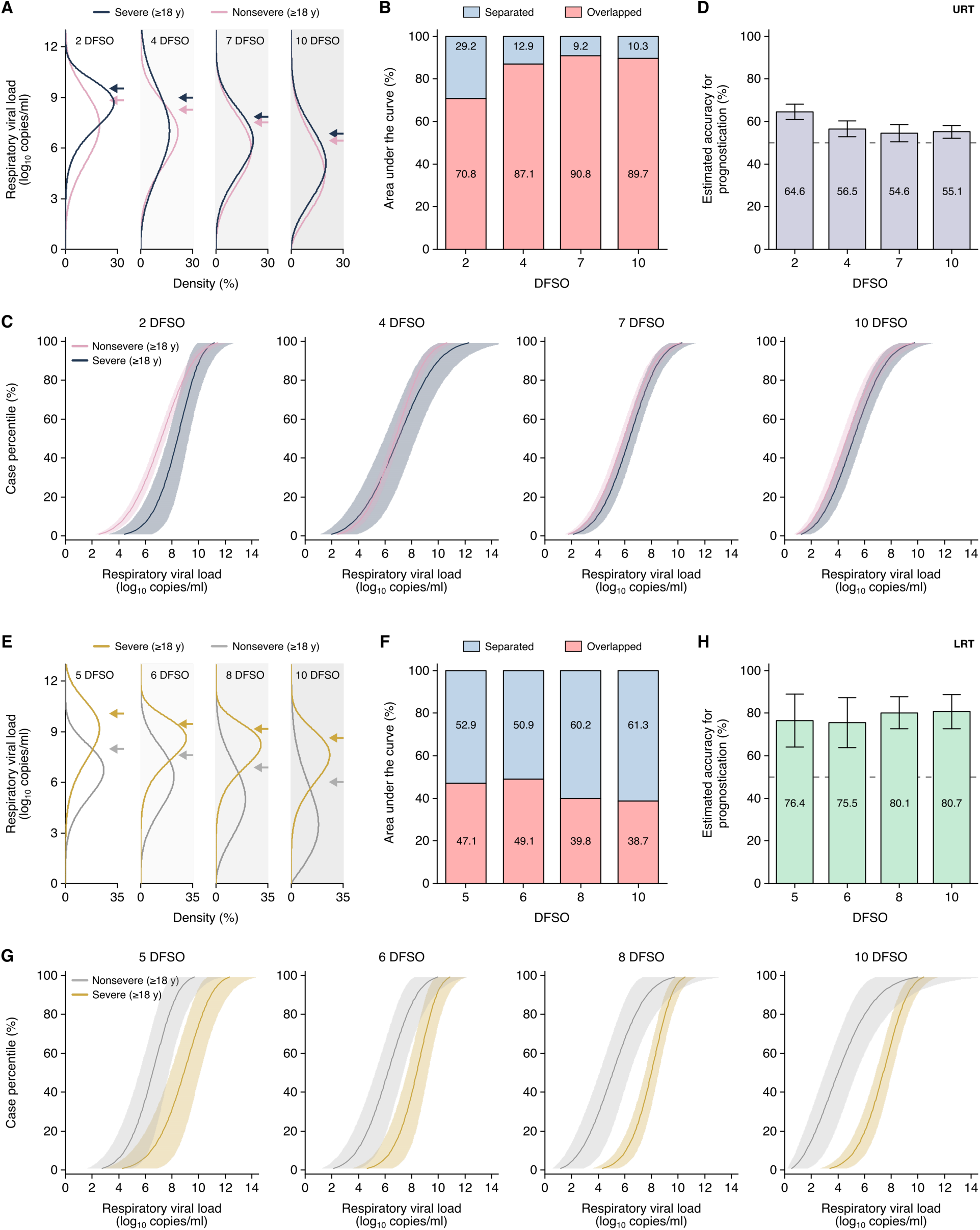
**Heterogeneity in, and severity prognostication from, SARS-CoV-2 shedding for adult COVID-19**. (**A**) Estimated distributions at 2, 4, 7 and 10 days from symptom onset (DFSO) of upper respiratory tract (URT) shedding for adults (aged 18 y or older) with nonsevere or severe COVID-19. (**B**) Overlapped or separated areas under the curve for the distributions in (A). (**C**) Cumulative distributions of URT shedding for adults with nonsevere or severe COVID-19 at various DFSO. (**D**) Estimated accuracy for using URT shedding of SARS-CoV-2 as a prognostic indicator for COVID-19 severity. (**E**) Estimated distributions at 5, 6, 8 and 10 DFSO of lower respiratory tract (LRT) shedding for adults with nonsevere or severe COVID-19. (**F**) Overlapped or separated areas under the curve for the distributions in (E). (**G**) Cumulative distributions of LRT shedding for adults with nonsevere or severe COVID-19 at various DFSO. (**H**) Estimated accuracy for using LRT shedding of SARS-CoV-2 as a prognostic indicator for COVID-19 severity. Arrows in (A) and (E) denote the 80^th^ case percentiles, in terms of rVL, for each group. For (C) and (G), the proportion of cases to the left of a given prognostic threshold are predicted to have nonsevere COVID-19 while those to the right of it are predicted to have severe disease. Sensitivity and specificity can then be estimated using the nonsevere and severe distributions. The dotted lines in (D) and (H) denote 50% accuracy.

Since cases with severe COVID-19 tend to deteriorate at 10 DFSO (Berlin et al., 2020; Zhou et al., 2020), the early differences in shedding may predict disease severity. To assess the prognostic utility of URT shedding, we used the rVL distributions of nonsevere and severe adult cases and calculated the area under the curve (AUC) that is overlapped or separated (***Figure 4B***). The greater the separation between these rVL distributions, the greater the ability to differentiate severe COVID-19 from nonsevere illness, and this AUC analysis estimates the maximal accuracy of prognostication (Methods). At each DFSO, these URT distributions were largely overlapped. Moreover, the cumulative density distributions of rVL (***Figure 4C***) estimated poor sensitivity and specificity for prognostication (***Figure 4—Figure Supplement 2***). Thus, our data indicated that URT shedding inaccurately predicts COVID-19 severity (***Figure 4D***).

### Heterogeneity in LRT shedding of SARS-CoV-2

In contrast, the distributions of severe and nonsevere LRT shedding bifurcated along disease course (***Figure 4E***). At 6 DFSO, the estimate at the 80^th^ case percentile (cp) of LRT rVL was 9.40 (95% CI: 8.67-10.20) log_10_ copies/ml for severe COVID-19, while it was 7.66 (95% CI: 6.65-8.83) log_10_ copies/ml for nonsevere illness. At 10 DFSO, the difference between 80^th^-cp estimates expanded, as they were 8.63 (95% CI: 8.04-9.26) and 6.01 (95% CI: 4.65-7.78) log_10_ copies/ml for severe and nonsevere disease, respectively. Moreover, our data indicated that nonsevere illness was associated with greater skewing in LRT shedding than severe disease in the analyzed period (***Figure 4E***). For nonsevere COVID-19, the SD of rVL was 1.92, 2.01 and 2.09 log_10_ copies/ml at 6, 8 and 10 DFSO, respectively. For severe disease, it was lesser at 1.25, 1.37 and 1.61 log_10_ copies/ml for 6, 8 and 10 DFSO, respectively. On each DFSO, the top 2-12% of cases harbored 80% of the LRT copies of SARS-CoV-2 for adults with nonsevere COVID-19, whereas it was the top 10-20% of cases for adults with severe disease (***Figure 4—Figure Supplement 1, E to H***).

We also assessed the prognostic utility of LRT shedding. We calculated the area under the curve (AUC) that is overlapped or separated, which showed greater separation between the LRT distributions of severe and nonsevere cases (***Figure 4, F and G***). The estimated accuracy for using LRT shedding as a prognostic indicator for COVID-19 severity was up to 81% (***Figure 4H***). We used the cumulative distributions of LRT shedding (***Figure 4G***) to estimate the specificity and sensitivity at different prognostic thresholds of LRT shedding. For example, at 5 DFSO, the estimated specificity was 93.3% and the estimated sensitivity was 64.4% at a prognostic threshold of 9.10 log_10_ copies/ml (***Figure 4—Figure Supplement 3***). For 8 DFSO, the estimated specificity and sensitivity was 73.1 and 88.8%, respectively, at a prognostic threshold of 5.95 log_10_ copies/ml. These estimated specificities and sensitivities agreed with the estimated accuracy for prognostication. Taken together, our data indicated that LRT shedding more accurately predicts COVID-19 severity than does URT shedding.

## DISCUSSION

Our study systematically developed a dataset of COVID-19 case characteristics and rVLs and conducted stratified analyses on SARS-CoV-2 shedding post-symptom onset. In the URT, we found that adults with severe COVID-19 showed higher rVLs shortly after symptom onset, but similar SARS-CoV-2 clearance rates, when compared with their nonsevere counterparts. In the LRT, we found that high, persistent shedding was associated with severe COVID-19, but not nonsevere illness, in adults. Interestingly, in the analyzed periods, adults with severe disease tended to have higher rVLs in the LRT than the URT.

After stratifying for disease severity, we found that sex and age had nonsignificant effects on post-symptom-onset SARS-CoV-2 shedding levels and dynamics for each included analysis (summarized in ***Appendix Table 4***). Thus, while sex and age influence the tendency to develop severe COVID-19 (Onder et al., 2020; Tartof et al., 2020; Zhou et al., 2020), we find no such sex dimorphism or age distinction in shedding among cases of similar severity. This includes children, who had nonsevere illness in our study and show similar URT shedding post-symptom onset as adults with nonsevere illness.

Notably, our analyses indicate that high, persistent LRT shedding of SARS-CoV-2 characterizes severe COVID-19 in adults. This suggests that the effective immune responses associated with milder COVID-19, including innate, cross-reactive and coordinated adaptive immunity (Lucas et al., 2020; Ng et al., 2020; Pierce et al., 2020; Rydyznski et al., 2020; Takahashi et al., 2020), do not significantly inhibit early, or prolonged, SARS-CoV-2 replication in the LRT of severely affected adults. Hence, uncontrolled LRT replication tends to continue, at least, to 10 DFSO, coinciding with the timing of clinical deterioration (median, 10 DFSO) (Berlin et al., 2020; Zhou et al., 2020). Furthermore, the bifurcated profiles of LRT shedding concur with the observed severity-associated differences in lung pathology, in which severe cases show hyperinflammation and progressive loss of epithelial-endothelial integrity (Magro et al., 2020; Matheson & Lehner, 2020; Z. Xu et al., 2020).

Our results indicate that LRT shedding may be a prognostic indicator in SARS-CoV-2 infection, predicting disease severity before clinical deterioration. They reinforce that severe COVID-19 is associated with higher rVLs than nonsevere illness and suggest that sex and age may not significantly influence prognostic thresholds. Nonsevere and severe cases tend to clear SARS-CoV-2 from the LRT at different rates. Thus, time course of disease (e.g., DFSO) should be considered alongside rVL, rather than simply employing rVL at admission. Unlike URT shedding (our study predominantly analyzed nasopharyngeal and oropharyngeal swabs for URT specimens), LRT shedding bifurcates considerably between nonsevere and severe COVID-19, and SARS-CoV-2 quantitation from the LRT should more accurately predict severity. While URT specimens are typically used to diagnose COVID-19, LRT specimens (our study predominantly analyzed sputum) may be collected, especially from high-risk patients, for severity prognostication.

While our analyses did not account for virus infectivity, higher SARS-CoV-2 rVL is associated with a higher likelihood of culture positivity, from adults (van Kampen et al., 2021; Wolfel et al., 2020) as well as children (L’Huillier, Torriani, Pigny, Kaiser, & Eckerle, 2020), and higher transmission risk (Marks et al., 2021). Hence, our results suggest that infectiousness increases with COVID-19 severity, concurring with epidemiological analyses (Li et al., 2021; Sayampanathan et al., 2021). They also suggest that adult and pediatric infections of similar severity have comparable infectiousness, reflecting epidemiological findings on age-based infectiousness (Laxminarayan et al., 2020; Li et al., 2021; K. Sun et al., 2021). Moreover, since respiratory aerosols are typically produced from the LRT (Johnson et al., 2011), severe SARS-CoV-2 infections may have increased, and extended, risk for aerosol transmission. As severe cases tend to be hospitalized, this provides one possible explanation for the elevated risk of COVID-19 among healthcare workers in inpatient settings (Nguyen et al., 2020); airborne precautions, such as the use of N95 or air-purifying respirators, should be implemented around patients with COVID-19.

Our study has limitations. First, while our study design systematically developed a large, diverse dataset, there were few severe female cases with LRT specimens and no severe pediatric cases included. Statistical comparisons involving these groups were not conducted, and additional studies should permit these remaining comparisons. Second, our analyses did not assess the influence of additional case characteristics, including comorbidities, and their relationships with SARS-CoV-2 kinetics remain unclear. Third, the systematic dataset consisted largely of hospitalized patients, and our results may not generalize to asymptomatic infections.

In summary, our findings provide insight into SARS-CoV-2 kinetics and describe virological factors that mediate the clinical manifestations of COVID-19. They show that high, persistent LRT shedding characterizes severe disease in adults, highlighting the potential prognostic utility of SARS-CoV-2 quantitation from LRT specimens. Lastly, each study identified by our systematic review collected specimens before October 2020. As widespread transmission of the emerging variants of concerns likely occurred after this date (Davies et al., 2021; Tegally et al., 2021), our study presents a quantitative resource to assess the effects of their mutations on respiratory shedding levels and dynamics.

## METHODS

### Data sources and searches

Our systematic review identified studies reporting SARS-CoV-2 quantitation in respiratory specimens taken during the estimated infectious period (-3 to 10 days from symptom onset [DFSO]) (He et al., 2020; Wolfel et al., 2020). The systematic review protocol was based on our previous study (Chen et al., 2021) and was prospectively registered on PROSPERO (registration number, CRD42020204637). The systematic review was conducted according to Cochrane methods guidance (Higgins et al., 2019). Other than the title of this study, we have followed PRISMA reporting guidelines (Moher, Liberati, Tetzlaff, Altman, & Group, 2009).

Up to 20 November 2020, we searched, without the use of filters or language restrictions, the following sources: MEDLINE (Ovid, 1946 to 20 Nov 2020), EMBASE (Ovid, 1974 to 20 Nov 2020), Cochrane Central Register of Controlled Trials (via Ovid, 1991 to 20 Nov 2020), Web of Science Core Collection (up to 20 Nov 2020), and medRxiv and bioRxiv (both searched through Google Scholar via the Publish or Perish program, up to 20 Nov 2020). We also gathered studies by searching through the reference lists of review articles identified by the database search, by searching through the reference lists of included articles, through expert recommendation (by Epic J. Topol and Akiko Iwasaki on Twitter) and by hand-searching through journals. A comprehensive search was developed by a librarian (Z.P.). The line-by-line search strategies for all databases are included in ***Figure 1—Source Data 1 to 5***. The search results were exported from each database and uploaded to the Covidence online system for deduplication and screening.

### Study selection

Studies that reported SARS-CoV-2 quantitation in individual URT (nasopharyngeal swab [NPS], nasopharyngeal aspirate [NPA], oropharyngeal swab [OPS] or posterior oropharyngeal saliva [POS]) or LRT (endotracheal aspirate [ETA] or sputum [Spu]) specimens taken during the estimated infectious period (-3 to 10 DFSO) in humans were included (additional details in the **Appendix**). As semiquantitative metrics (cycle threshold [Ct] values) cannot be compared on an absolute scale between studies based on instrument and batch variation (Han, Byun, Cho, & Rim, 2021), studies reporting specimen measurements as Ct values, without quantitative calibration, were excluded. Two authors (P.Z.C. and N.B.) independently screened titles and abstracts and reviewed full texts. At the full-text stage, reference lists were reviewed for study inclusion. Inconsistencies were resolved by discussion and consensus, and 26 studies met the inclusion criteria (Bal et al., 2020; Benotmane et al., 2020; Biguenet et al., 2021; Fajnzylber et al., 2020; Han et al., 2020; Hirotsu et al., 2020; Hurst et al., 2020; Iwasaki et al., 2020; L’Huillier et al., 2020; Lavezzo et al., 2020; Pan, Zhang, Yang, Poon, & Wang, 2020; Peng et al., 2020; Shrestha et al., 2020; J. Sun et al., 2020; To et al., 2020; van Kampen et al., 2021; Vetter et al., 2020; Wolfel et al., 2020; Wyllie et al., 2020; Y. Xu et al., 2020; Yazdanpanah, French, & study, 2021; Yilmaz et al., 2021; Yonker et al., 2020; Zhang et al., 2020; Zheng et al., 2020; Zou et al., 2020). Additional details on study selection can be found in our previous protocol (Chen et al., 2021)

### Data extraction and risk-of-bias assessment

Two authors (P.Z.C. and N.B.) independently collected data (specimen measurements taken between -3 and 10 DFSO, specimen type, volume of transport media and case characteristics, including age, sex and disease severity) from contributing studies and assessed risk of bias using a modified version of the Joanna Briggs Institute (JBI) tools for case series, analytical cross-sectional studies and prevalence studies (Moola et al., 2020; Munn et al., 2019; Munn, Moola, Lisy, Riitano, & Tufanaru, 2015) (shown in the **Appendix**). Items were judged with responses to data inquiries, if authors responded.

Data were collected for individually reported specimens of known type, with known DFSO, and for COVID-19 cases with known age, sex or severity. Case characteristics were collected directly from contributing studies when reported individually or obtained via data request from the authors. Data from serially sampled asymptomatic cases were included, and the day of laboratory diagnosis was referenced as 0 DFSO (Lavezzo et al., 2020; Wolfel et al., 2020). Based on the modified JBI checklist, studies were considered to have low risk of bias if they met the majority of items and included item 1 (representative sample). Discrepancies were resolved by discussion and consensus. Studies at high or unclear risk of bias typically included samples that were not representative of the target population; did not report the VTM volume used; had non-consecutive inclusion for case series and cohort studies or did not use probability-based sampling for cross-sectional studies; and did not report the response rate.

### Respiratory viral load

To enable analyses based on rVL (viral RNA concentration in the respiratory tract) and to account for interstudy variation in specimen measurements, the rVL for each collected sample was estimated based on the specimen concentration (viral RNA concentration in the specimen) and its dilution factor in VTM. Typically, swabbed specimens (NPS and OPS) report the viral RNA concentration in VTM. Based on the VTM volume reported in the study along with the expected uptake volume for swabs (0.128 ± 0.031 ml, mean ± SD) (Warnke, Warning, & Podbielski, 2014), we calculated the dilution factor for each respiratory specimen and then estimated the rVL. Similarly, liquid specimens (ETA, POS and Spu) are often diluted in VTM, and the rVL was estimated based on the reported collection and VTM volumes. If the diluent volume was not reported, then VTM volumes of 1 ml (NPS and OPS) or 2 ml (POS and ETA) were assumed (Lavezzo et al., 2020; To et al., 2020). Unless dilution was reported, Spu specimens were taken as undiluted (Wolfel et al., 2020). The non-reporting of VTM volume was noted as an element increasing risk of bias in the modified JBI critical appraisal checklist. For laboratory-confirmed COVID-19 cases, negative specimen measurements were taken at the reported assay detection limit in the respective study.

### Case definitions

As severity in the clinical manifestations of COVID-19 and case-fatality rates tend to increase among children (aged 0-17 y), younger adults (aged 18-59 y) and older adults (aged 60 y or older) (Onder et al., 2020; Zheng et al., 2020), the data were delineated based on these three age groups. Cases were also categorized by sex.

U.S. National Institutes of Health guidance was used to categorize disease severity as nonsevere or severe (Health). The nonsevere group included those with asymptomatic infection (individuals who test positive via a molecular test for SARS-CoV-2 and report no symptoms consistent with COVID-19); mild illness (individuals who report any signs or symptoms of COVID-19, including fever, cough, sore throat, malaise, headache, muscle pain, nausea, vomiting, diarrhea, loss of taste and smell, but who do not have dyspnea or abnormal chest imaging); and moderate illness (individuals with clinical or radiographic evidence of LRT disease, fever >39.4°C or SpO_2_ ≥94% on room air) disease. The severe group included those with severe illness (individuals who have SpO_2_ <94% on room air, [PaO_2_/FiO_2_] <300 mmHg, respiratory rate >30 breaths/min or lung infiltrates >50%) and critical illness (respiratory failure, septic shock or multiple organ dysfunction).

### Regression analyses

We used regression analysis to assess the respiratory shedding of SARS-CoV-2 and compare age, sex or severity groups. In COVID-19 cases, rVL tends to diminish exponentially after 1 DFSO in the URT, whereas it tends to do so after 4 DFSO in the LRT (Bernheim et al., 2020; Chen et al., 2021; Wolfel et al., 2020). Hence, rVLs (in units of log_10_ copies/ml) between 1-10 DFSO for the URT, or 4-10 DFSO for the LRT, were fitted using general linear regression with interaction:

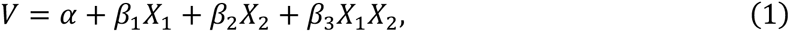

where *V* represents the rVL, α represents the estimated mean rVL (at 1 DFSO for URT or 4 DFSO for LRT) for the reference group, *X*_1_ represents DFSO for the reference group, *X*_2_ represents the comparison group, *β*_1_ represents the effect of DFSO on rVL for the reference group, *β*_2_ represents the effect of the comparison group on the intercept and *β*_3_ represents the interaction between DFSO and groups. Regression analyses were offset by DFSO such that mean rVLs at 1 DFSO for URT, or 4 DFSO for LRT, were compared between groups by the effect on the intercept (regression *t*-test for *β*_3_). Shedding dynamics were compared between groups by interaction (regression *t*-test for *β*_3_). The statistical significance of viral clearance for each group was analyzed using simple linear regression (regression *t*-test on the slope). Regression models were extrapolated (to 0 log_10_ copies/ml, rather than an assay detection limit) to estimate the duration of shedding. Each group in statistical analyses included all rVLs for which the relevant characteristic (LRT or URT, age group, sex or disease severity) was ascertained at the individual level. Groups with small sample sizes were not compared, as these analyses are more sensitive to potential sampling error. Statistical analyses were performed using OriginPro 2019b (OriginLab) and the General Linear regression app. *P* values below 0.05 were considered statistically significant.

### Distribution analyses

To assess heterogeneity in shedding, rVL data were fitted to Weibull distributions (Chen et al., 2021). The Weibull quantile function and Weibull cumulative distribution function were used to estimate the rVL at a case percentile and the percentage of cases at a given rVL, respectively. Each distribution was fitted to groups that included all rVLs for which the relevant characteristic (LRT or URT, age group, sex or disease severity) was ascertained at the individual level. Distribution fitting was performed using Matlab R2019b (MathWorks) and the Distribution Fitter app.

### Prognostication accuracy

The fitted Weibull distributions were used to estimate the accuracy when using URT or LRT rVLs of SARS-CoV-2 as a prognostic indicator for SARS-CoV-2 infection. The overlapped AUC and separated AUC were calculated using the rVL distributions for severe and nonsevere adult COVID-19. These calculations were performed on separate DFSO and, separately, for the URT and LRT. The estimated maximal accuracy for prognostication at a given rVL threshold was then estimated by *A* = 50% + (50% ∗ *AUC_separated_*), where *AUC_separated_* represents the separated AUC for the nonsevere and severe distributions. The 95% CIs for prognostication accuracy were estimated using the proportional 95% CIs in the respective Weibull cumulative distributions. As the Weibull cumulative distributions estimate the percentage of cases at a given rVL, they were also used to estimate the sensitivity and specificity at a given prognostic threshold of rVL. By definition, the cases with rVL lower than the prognostic threshold are predicted to have nonsevere COVID-19, whereas those with rVL above it are predicted to have severe COVID-19. Hence, we used the cumulative distributions for nonsevere and severe adult cases on a DFSO and calculated the proportion of cases that were true positive, false positive, false negative and true negative rates across prognostic thresholds of rVL. Sensitivity and specificity were calculated based on these values.

## Data Availability

Study protocol, statistical code and data set: Available from Dr. Gu.

## Acknowledgement

The authors thank S. Fafi-Kremer, PharmD, PhD (Strasbourg University Hospital); Y. Hirotsu, PhD (Yamanashi Central Hospital); M.S. Kelly, MD, MPH (Duke University); E. Lavezzo, PhD, and A. Crisanti, MD, PhD (University of Padova); J.Z. Li, MD, MMSc (Brigham & Women’s Hospital); Cédric Laouénan, MD, PhD, and Yazdan Yazdanpanah, MD, PhD (Bichat-Claude Bernard University Hospital); N.K. Shrestha, MD (Cleveland Clinic); T. Teshima, MD, PhD (Hokkaido University); S. Trouillet-Assant, PhD (Université Hospital of Lyon); J.J.A. van Kampen, MD, PhD (Erasmus University Medical Center); A. Wyllie, PhD, N. Grubaugh, PhD, and A. Ko, MD (Yale School of Public Health); and A. Yilmaz, MD, PhD (Sahlgrenska University Hospital) for responses to data inquiries.

## Financial support

This study was supported by NSERC. P.Z.C. was supported by the NSERC Vanier Canada Graduate Scholarship (608544). D.N.F was supported by the Canadian Institutes of Health Research (Canadian COVID-19 Rapid Research Fund, OV4-170360). F.X.G. was supported by the NSERC Discovery Grant, NSERC Senior Industrial Research Chair program and Toronto COVID-19 Action Fund.

## Data availability

The systematic dataset and model outputs from this study will be uploaded to Zenodo after peer review. The systematic review protocol was prospectively registered on PROSPERO (registration number, CRD42020204637).

## FIGURE SUPPLEMENTS

**Figure 1—Figure supplement 1.**
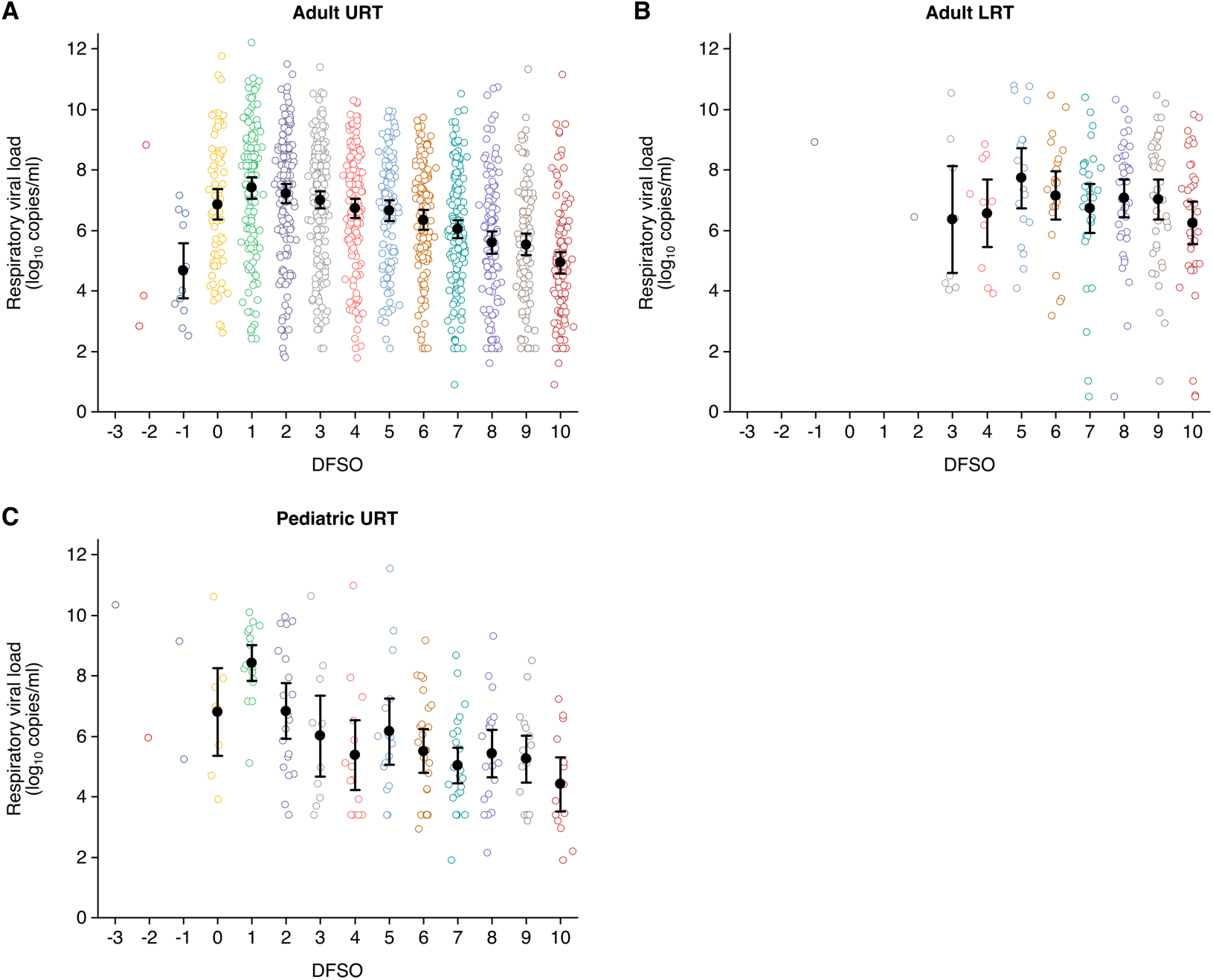
Summary of rVLs in the systematic dataset. (A) URT rVLs for adult COVID-19 cases (from -3 to 10 days from symptom onset [DFSO], *n* = 0, 3, 12, 71, 145, 159, 176, 152, 114, 123, 161, 125, 110, and 117 samples per DFSO). (B) LRT rVLs for adult COVID-19 cases (from -3 to 10 DFSO, *n* = 0, 0, 1, 0, 0, 1, 8, 10, 18, 23, 31, 39, 40, and 40 samples per DFSO). (C) URT rVLs for pediatric COVID-19 cases (from -3 to 10 DFSO, *n* = 1, 1, 2, 8, 17, 20, 11, 14, 16, 25, 26, 20, 17, and 14 samples per DFSO). Data were collected within the estimated infectious period of SARS-CoV-2 (-3 to 10 DFSO). The systematic search found no quantitative specimen measurements from the LRT for pediatric COVID-19. Open circles show rVL data. Filled circles and bars depict mean estimates and 95% CIs, respectively.

**Figure 2—Figure supplement 1.**
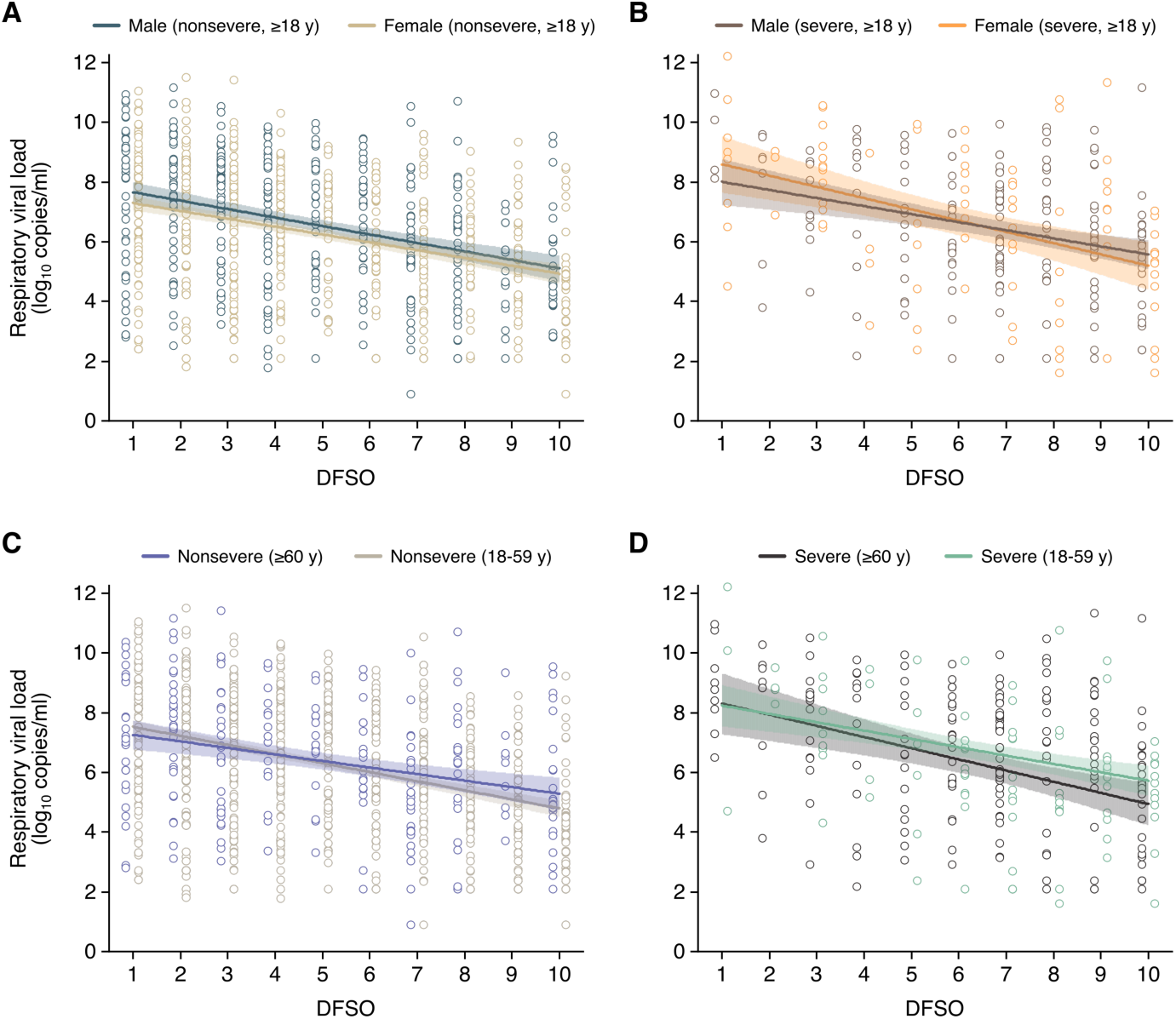
**SARS-CoV-2 shedding in the URT for adult COVID-19.** (**A** to **D**) Upper respiratory tract (URT) shedding for male and female adult (aged 18 y or older) cases with nonsevere COVID-19 (A), for male and female adult cases with severe COVID-19 (B), for older (aged 60 y or older) and younger (aged 18-59 y) adults with nonsevere COVID-19 (C) and for older and younger adults with severe COVID-19 (D). Open circles represent rVL data and were offset from their DFSO for visualization. Lines and bands show regressions and their 95% CIs, respectively.

**Figure 2—Figure supplement 2.**
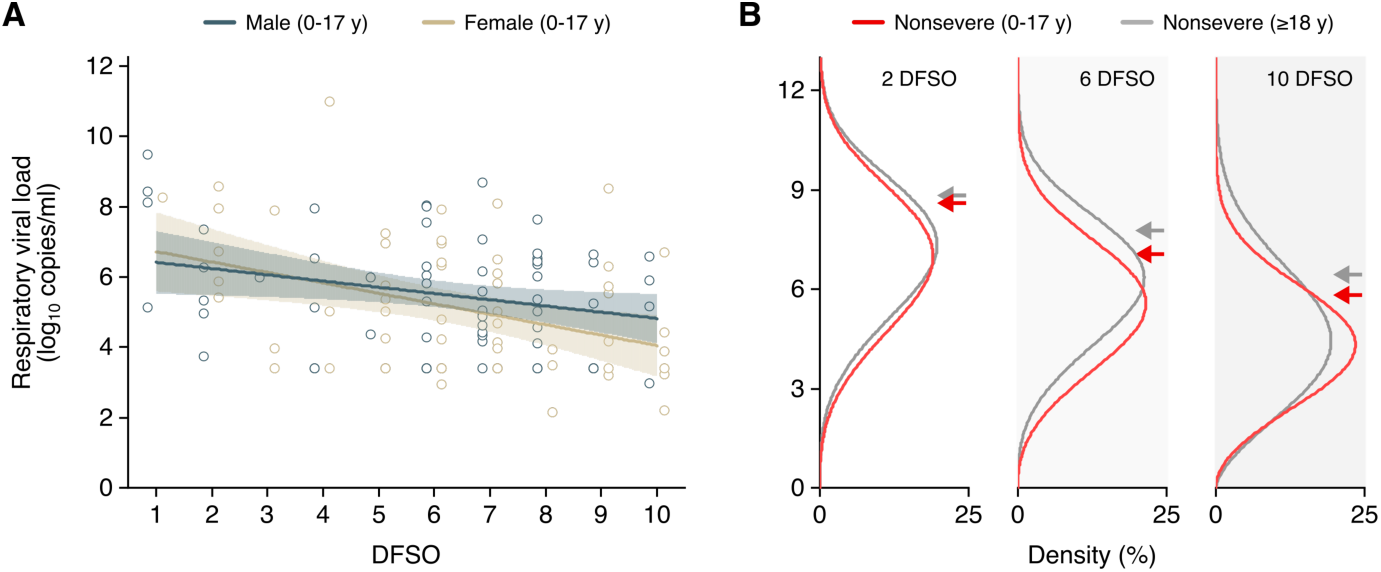
**SARS-CoV-2 shedding in the URT for pediatric COVID-20.** (**A**) Upper respiratory tract (URT) shedding for male and female pediatric (aged 0-17 y) cases with nonsevere COVID-19. Open circles represent rVL data and were offset from their DFSO for visualization. Lines and bands show regressions and their 95% CIs, respectively. (**B**) Estimated distributions at 2, 6 and 10 days from symptom onset (DFSO) of URT shedding for adult (aged 18 y or older) and pediatric cases with nonsevere COVID-19.

**Figure 3—Figure supplement 1.**
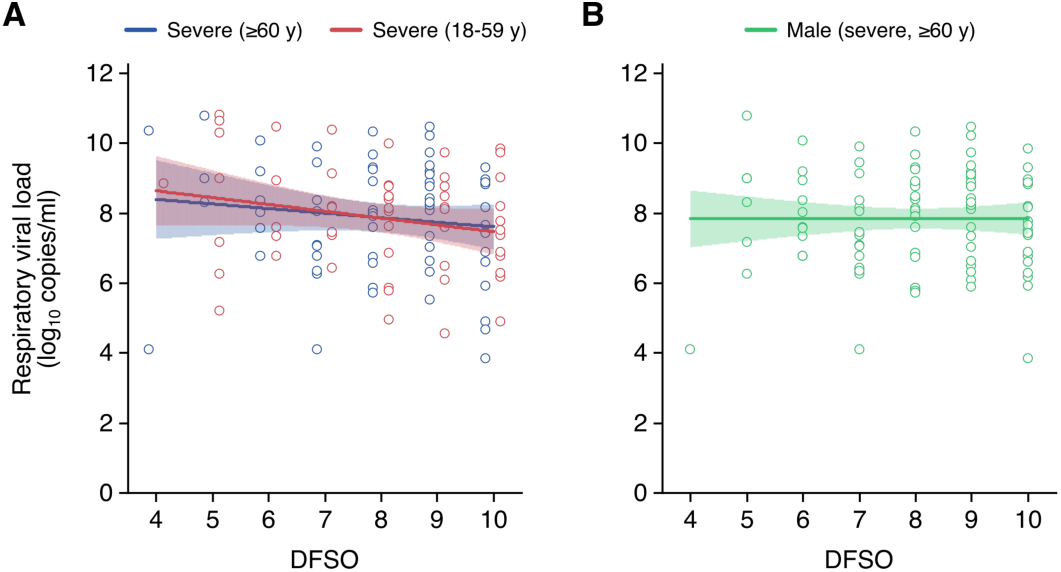
**SARS-CoV-2 shedding in the LRT for adult COVID-19.** (**A** and **B**) Lower respiratory tract (LRT) shedding for older (aged 60 y or older) or younger (aged 18-59 y) adult cases with severe COVID-19 (A) and male adult (aged 18 y or older) cases severe COVID-19 (B). Open circles represent rVL data and were offset from their DFSO for visualization. Lines and bands show regressions and their 95% CIs, respectively.

**Figure 4—Figure supplement 1.**
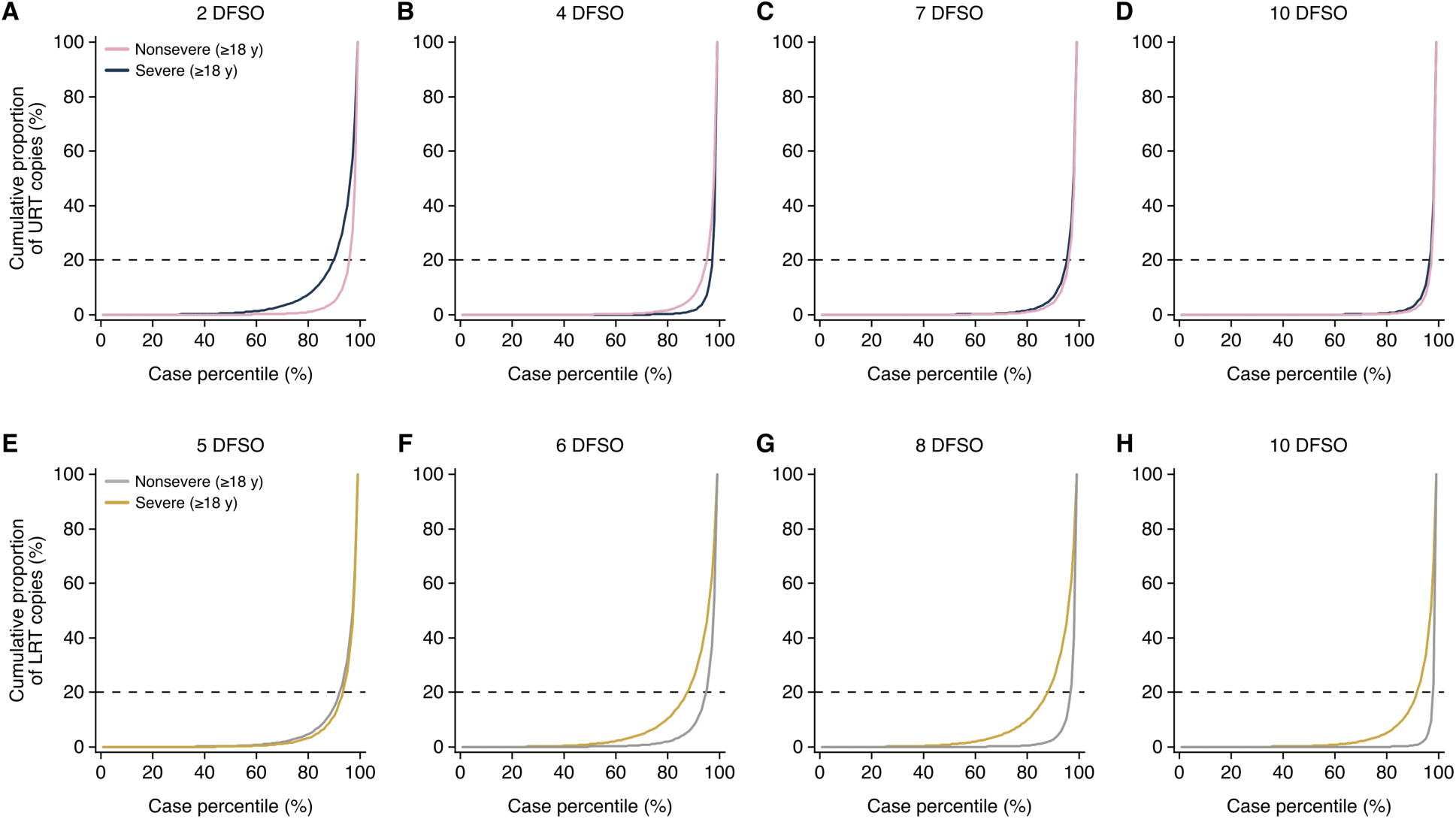
**Few cases carry the majority of SARS-CoV-2 copies in the URT and LRT.** (**A** to **D**) Cumulative distribution of upper respiratory tract (URT) SARS-CoV-2 copies harbored in adults with nonsevere or severe COVID-19 at 2 (A), 4 (B), 7 (C) and 10 (D) days from symptom onset (DFSO). (**E** to **H**) Cumulative distribution of lower respiratory tract (LRT) SARS-CoV-2 copies harbored in adults with nonsevere or severe COVID-19 at 5 (A), 6 (B), 8 (C) and 10 (D) DFSO. These curves were based on the fitted Weibull distributions in copies/ml and estimate the proportion of cases that carry the total amount of SARS-CoV-2 in the URT or LRT. The dotted lines denote when the upper case percentiles harbor 80% of copies.

**Figure 4—Figure supplement 2.**
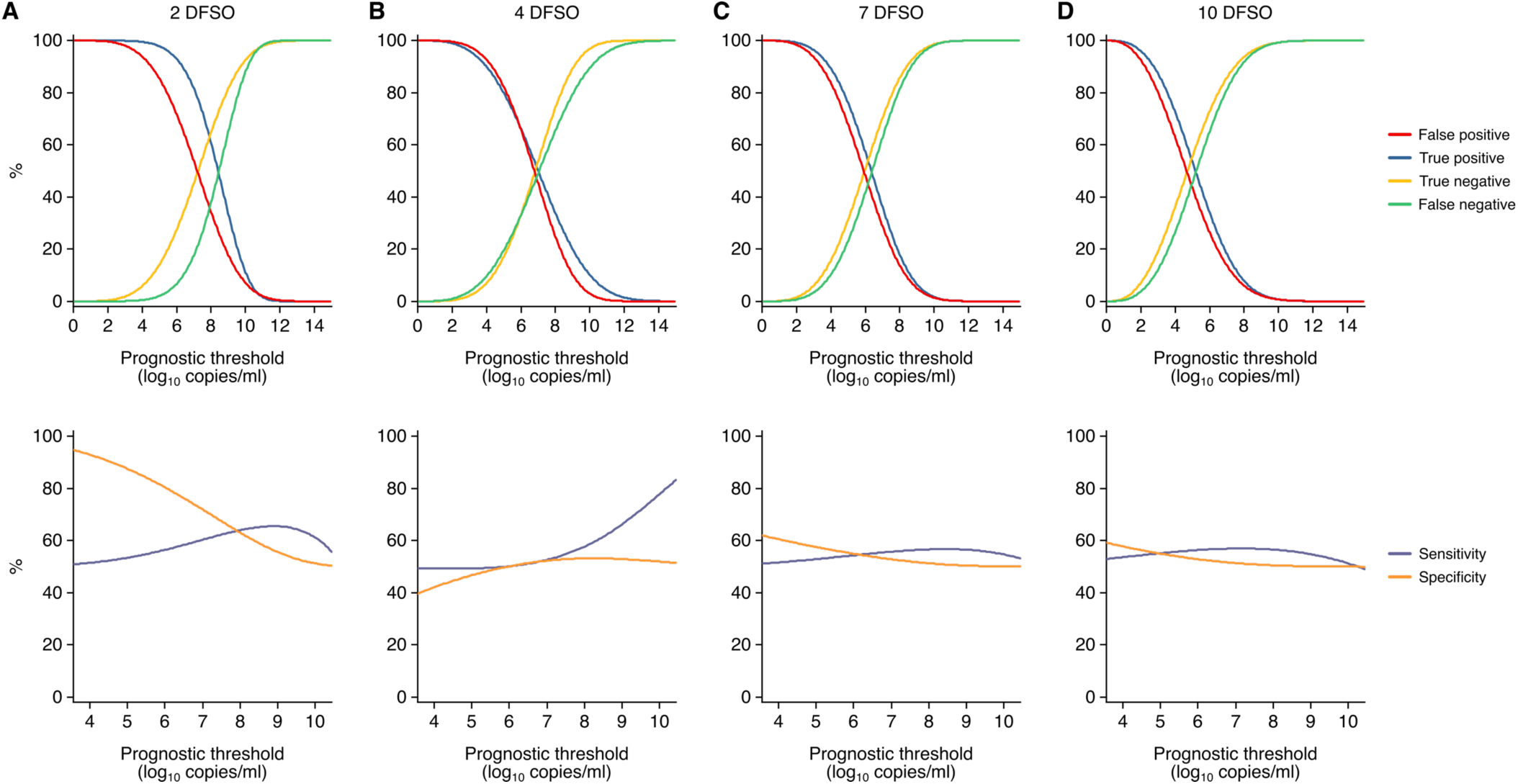
**Estimated sensitivity and specificity of URT shedding as a prognostic indicator for SARS-CoV-2 infection.** (**A** to **D**) Metrics for the accuracy of SARS-CoV-2 quantitation from the upper respiratory tract (URT) as a prognostic indicator for COVID-19 severity. Estimated true positive, true negative, false positive and false negative (top) or sensitivity and specificity (bottom) at 2 (A), 4 (B), 7 (C) and 10 (D) days from symptom onset (DFSO) across SARS-CoV-2 respiratory viral loads as the prognostic threshold. As depicted throughout this study, these plots show respiratory viral loads (rVLs, viral RNA concentration in the respiratory tract) rather than specimen concentrations (viral RNA concentration quantitated from a respiratory specimen).

**Figure 4—Figure supplement 3.**
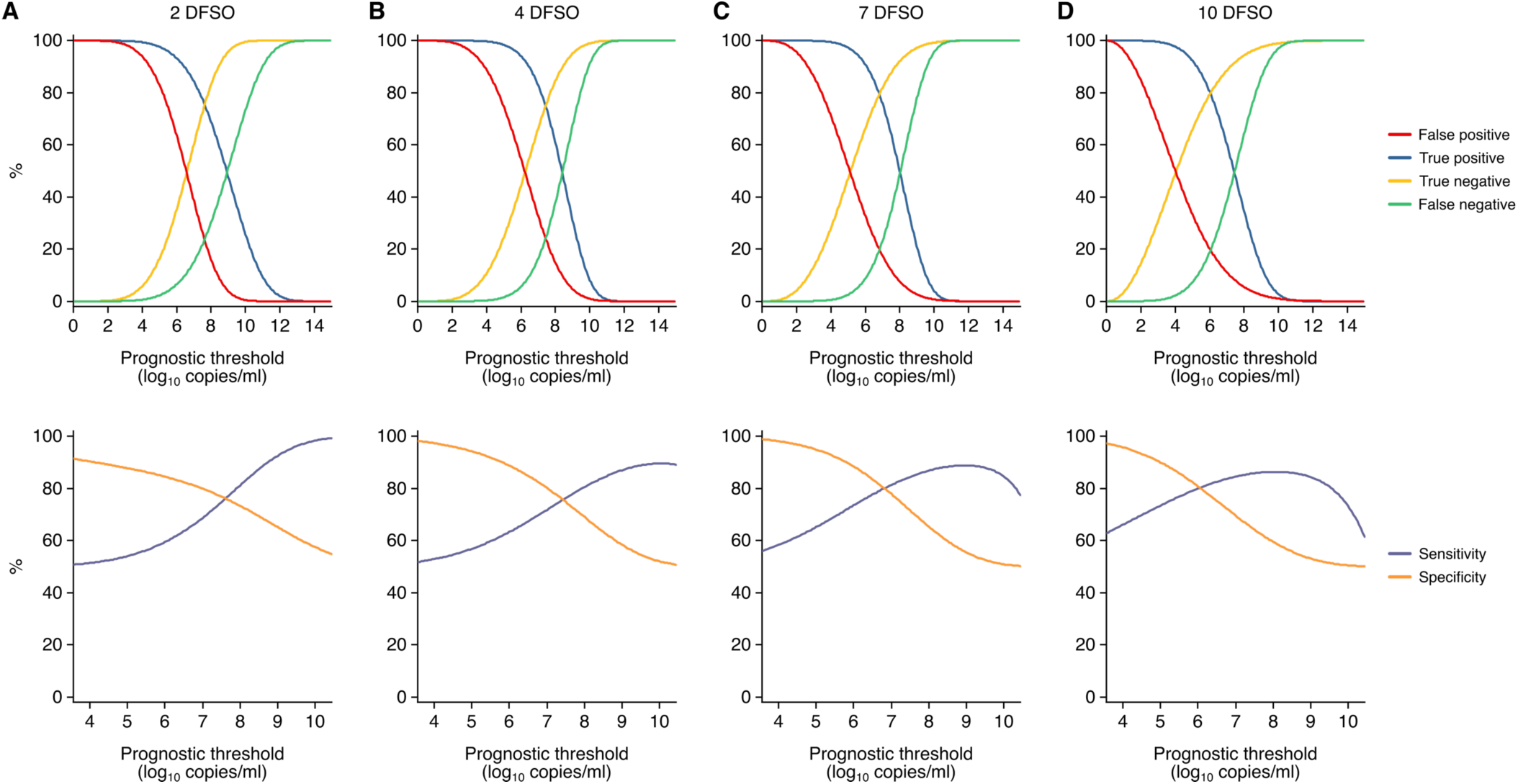
**Estimated sensitivity and specificity of LRT shedding as a prognostic indicator for SARS-CoV-2 infection.** (**A** to **D**) Metrics for the accuracy of SARS-CoV-2 quantitation from the lower respiratory tract (LRT) as a prognostic indicator for COVID-19 severity. Estimated true positive, true negative, false positive and false negative (top) or sensitivity and specificity (bottom) at 2 (A), 4 (B), 7 (C) and 10 (D) days from symptom onset (DFSO) across SARS-CoV-2 respiratory viral loads as the prognostic threshold. As depicted throughout this study, these plots show respiratory viral loads (rVLs, viral RNA concentration in the respiratory tract) rather than specimen concentrations (viral RNA concentration quantitated from a respiratory specimen).

## FIGURE DATA SOURCES

**Figure 1—Source Data 1.**
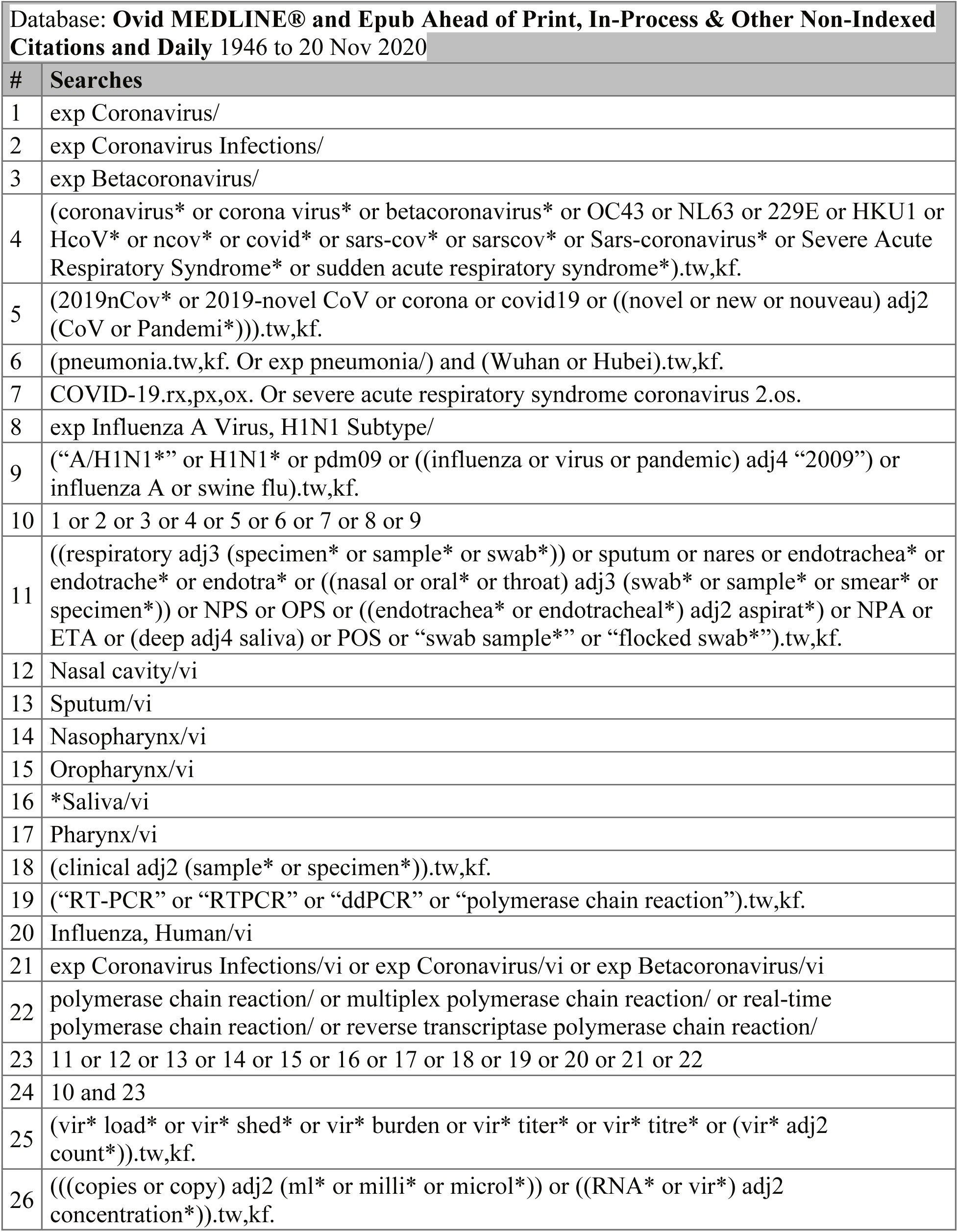
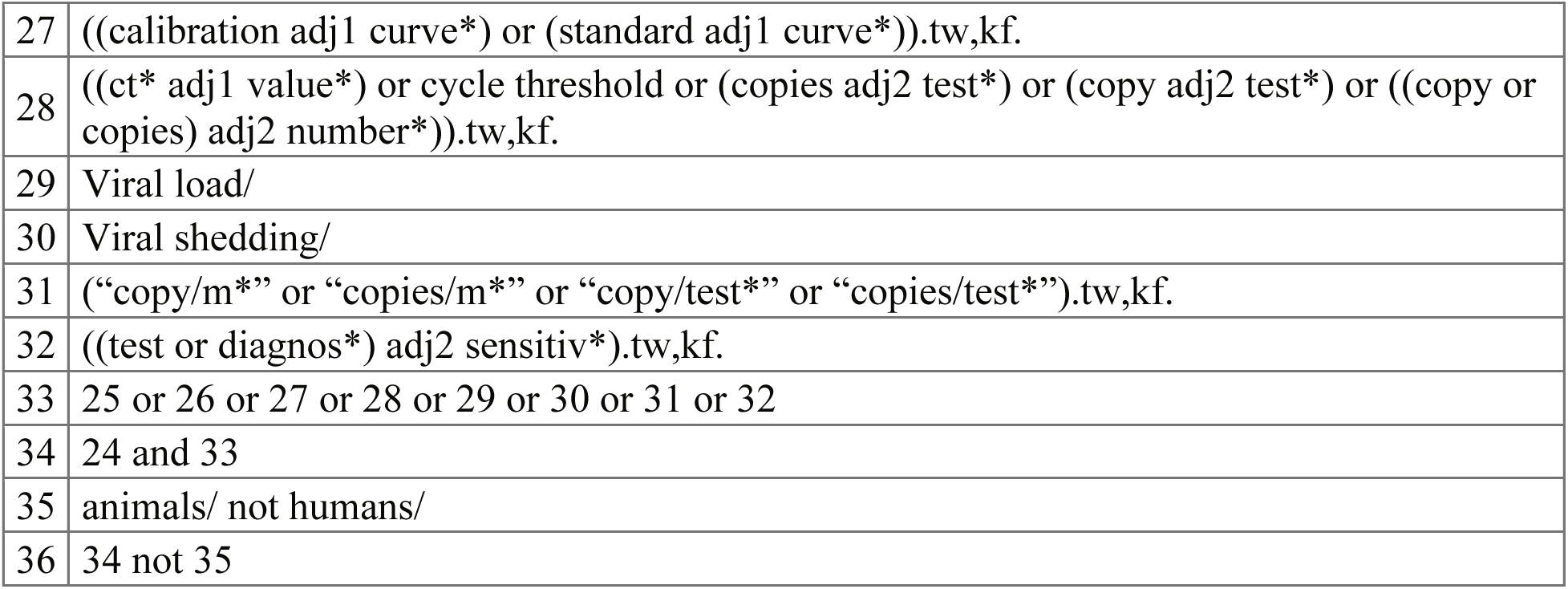
Search strategy used for MEDLINE.

**Figure 1— Source Data 2.**
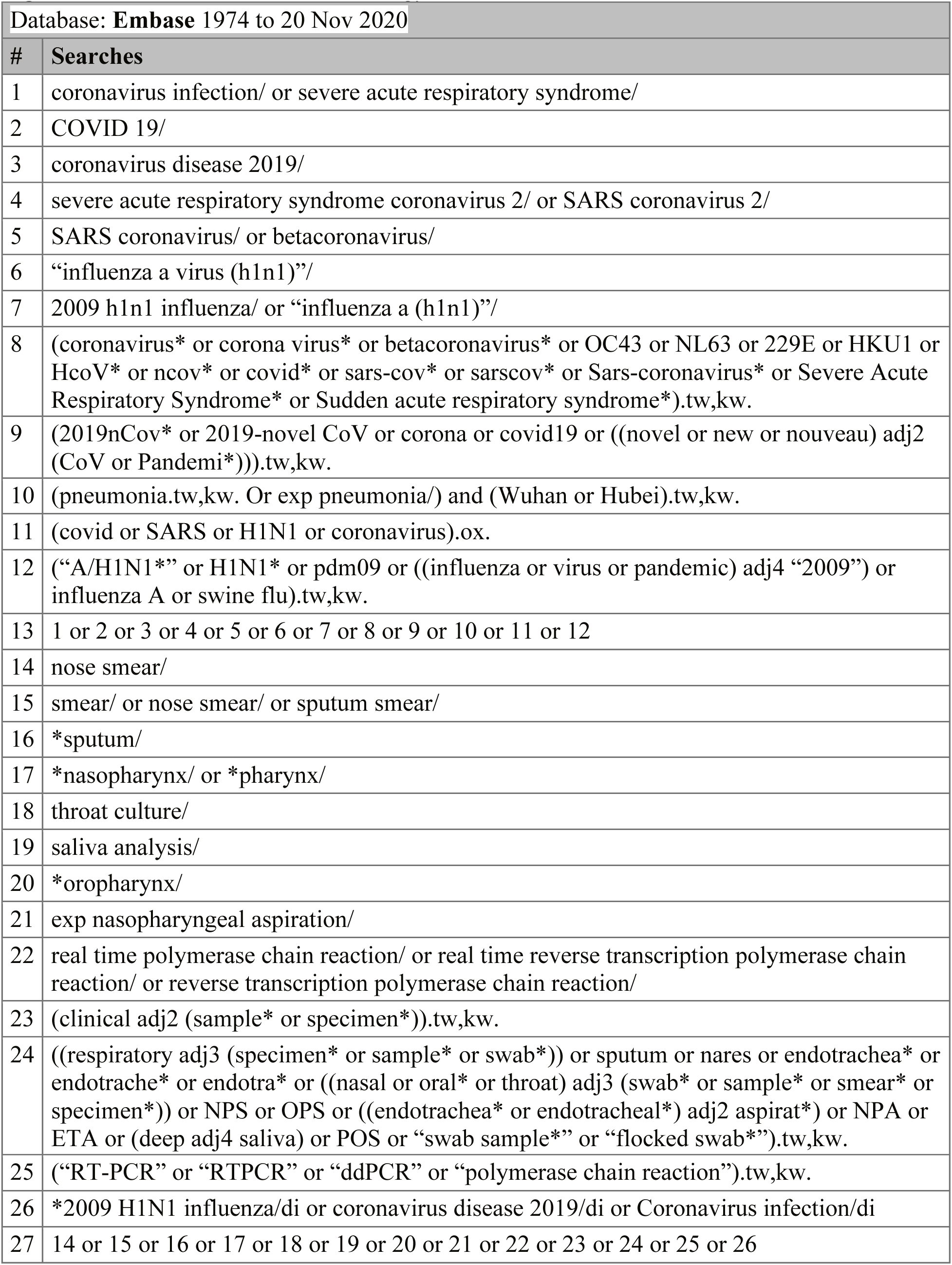
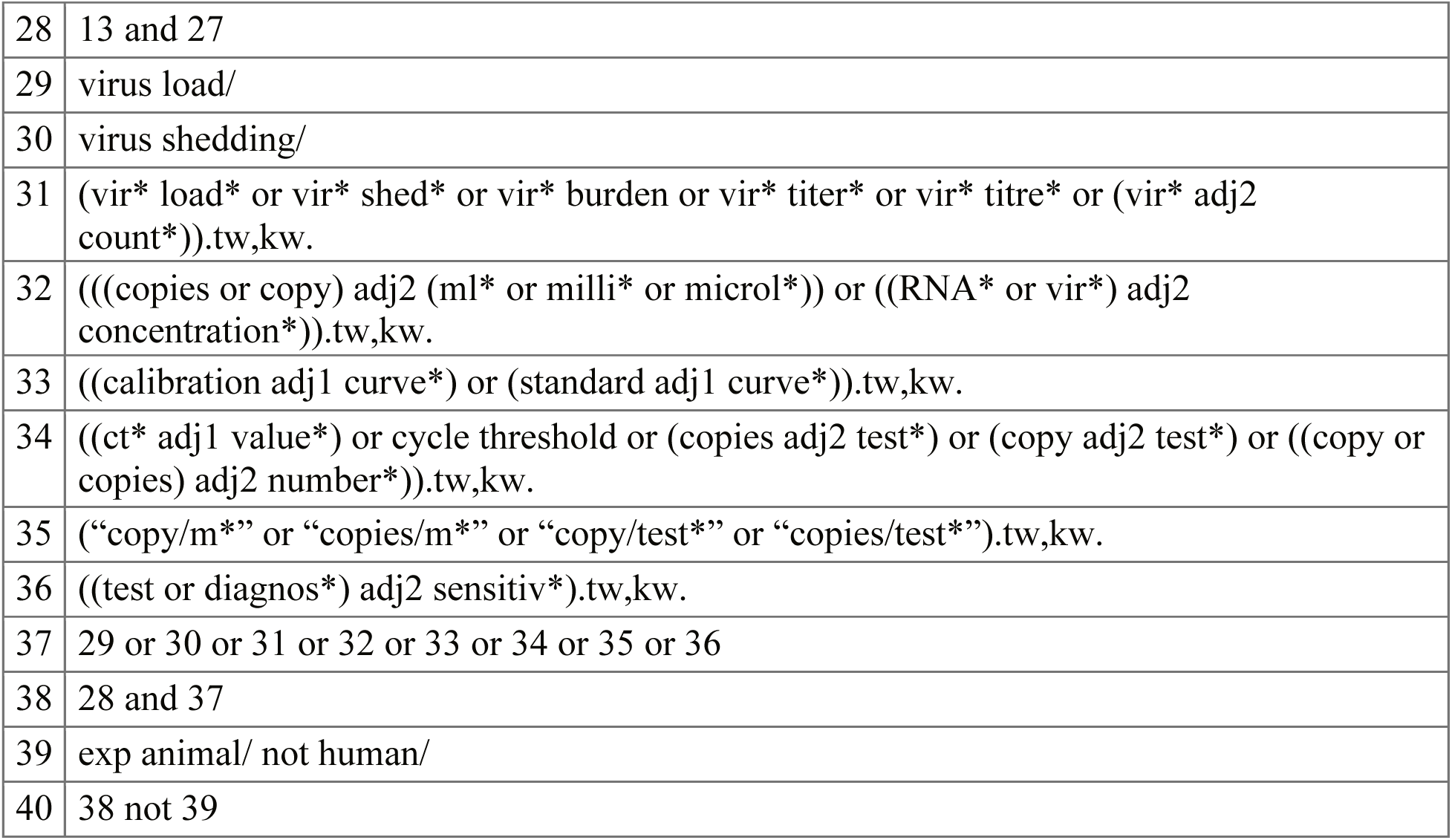
Search strategy used for EMBASE.

**Figure 1— Source Data 3.**
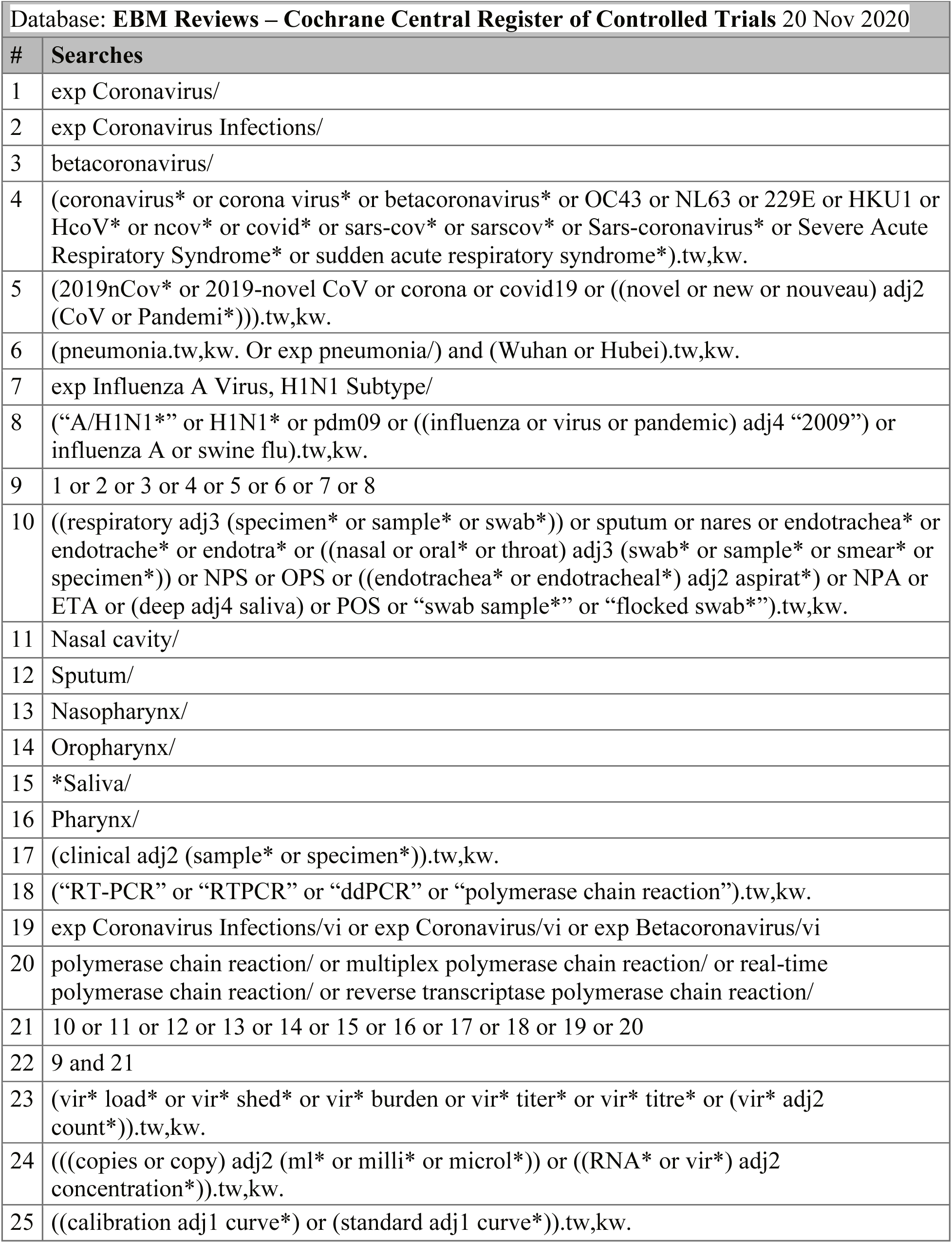
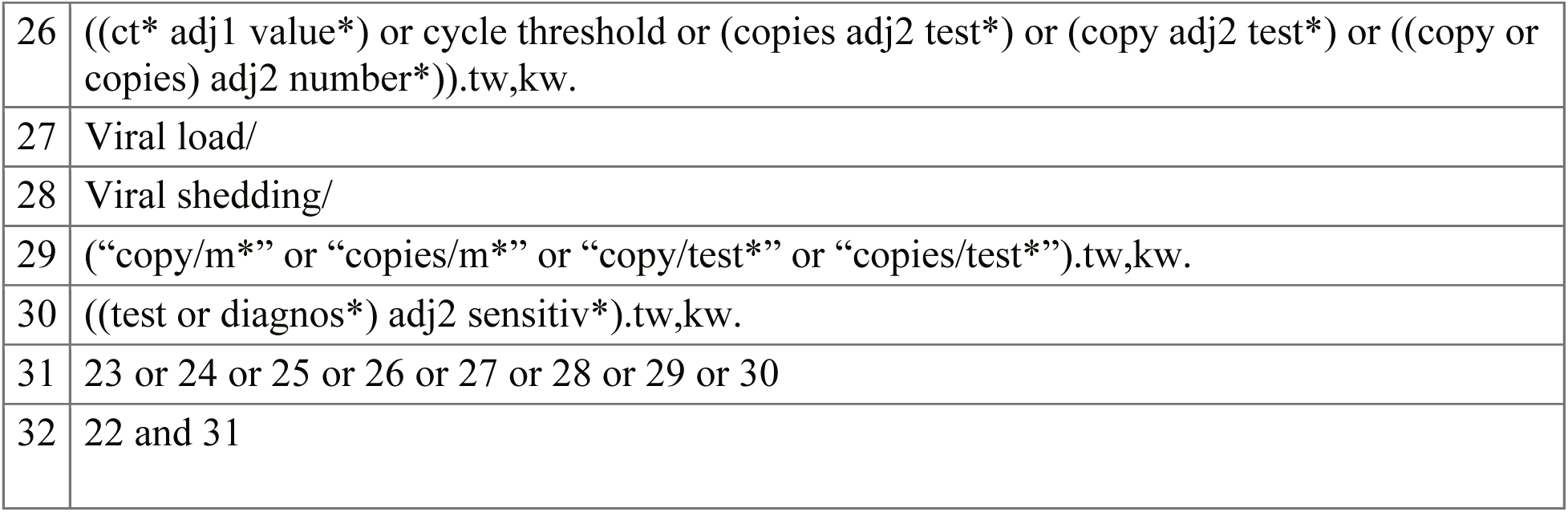
Search strategy used for Cochrane Central.

**Figure 1— Source Data 4.**
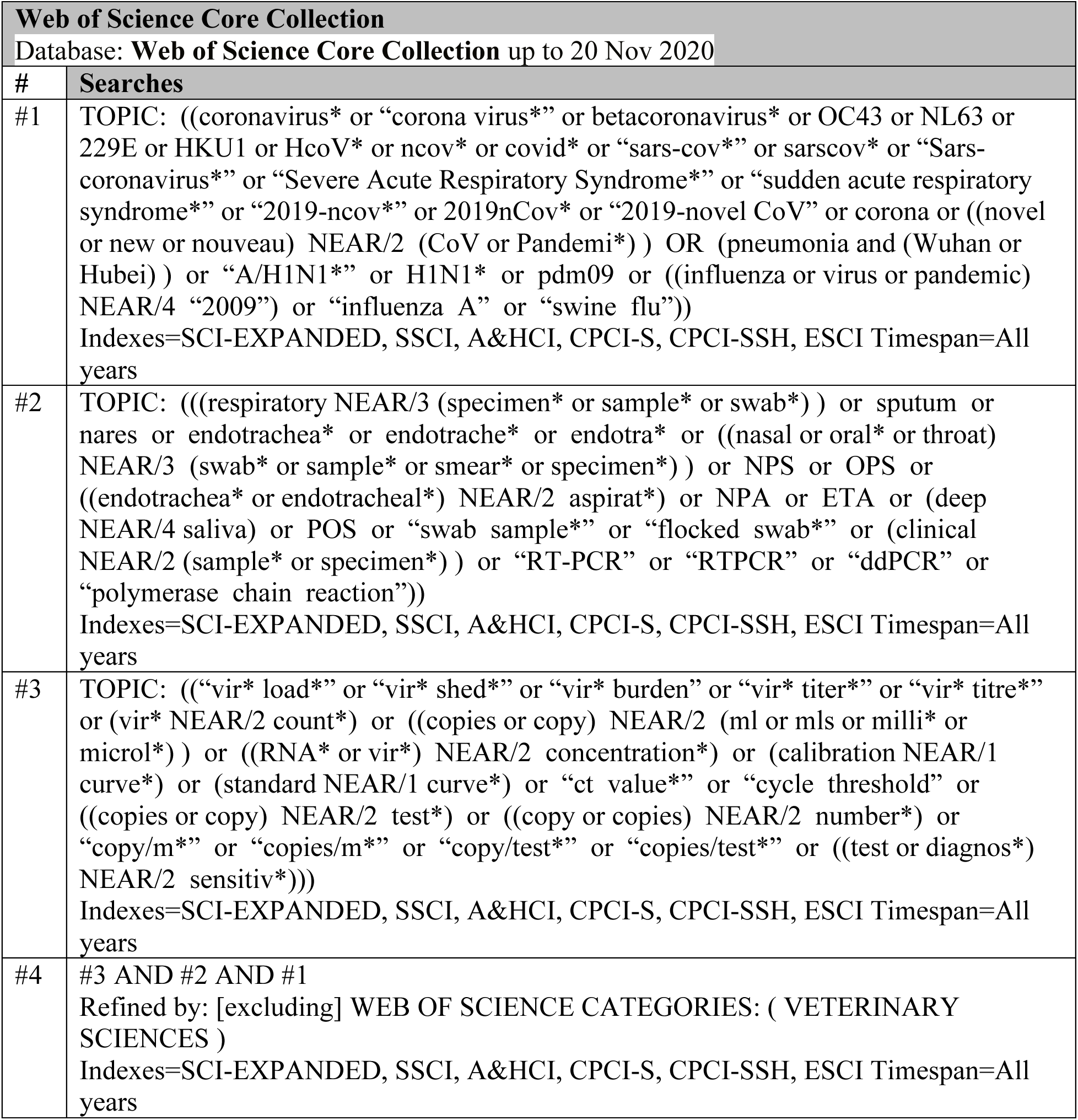
Search strategy used for Web of Science Core Collection.

**Figure 1—Source Data 5.**
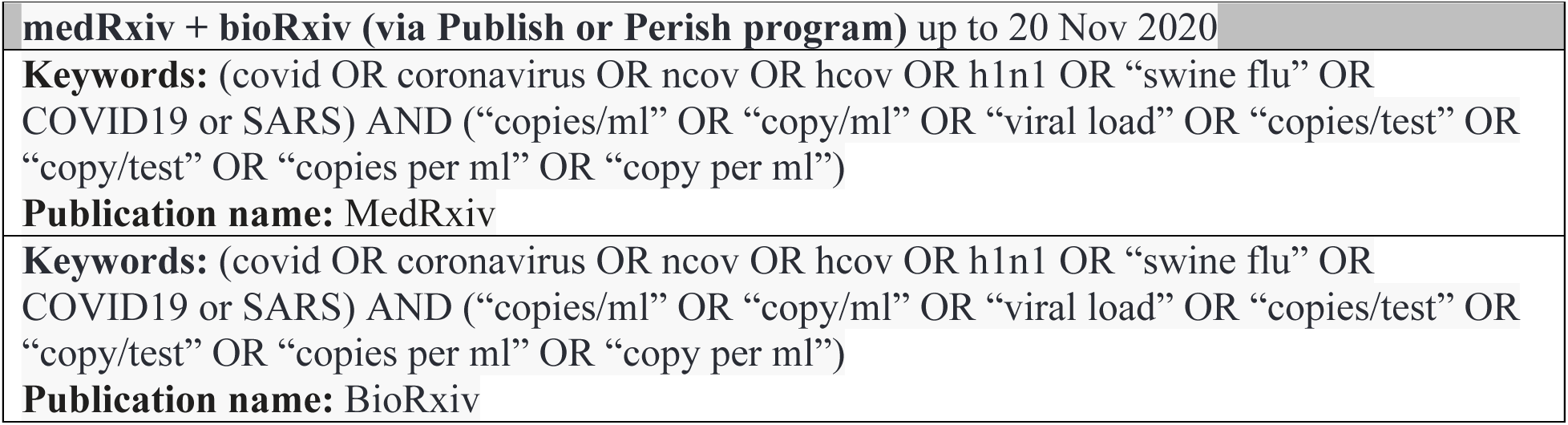
Search strategy used for medRxiv and bioRxiv.

## APPENDIX

**Appendix Table 1.**
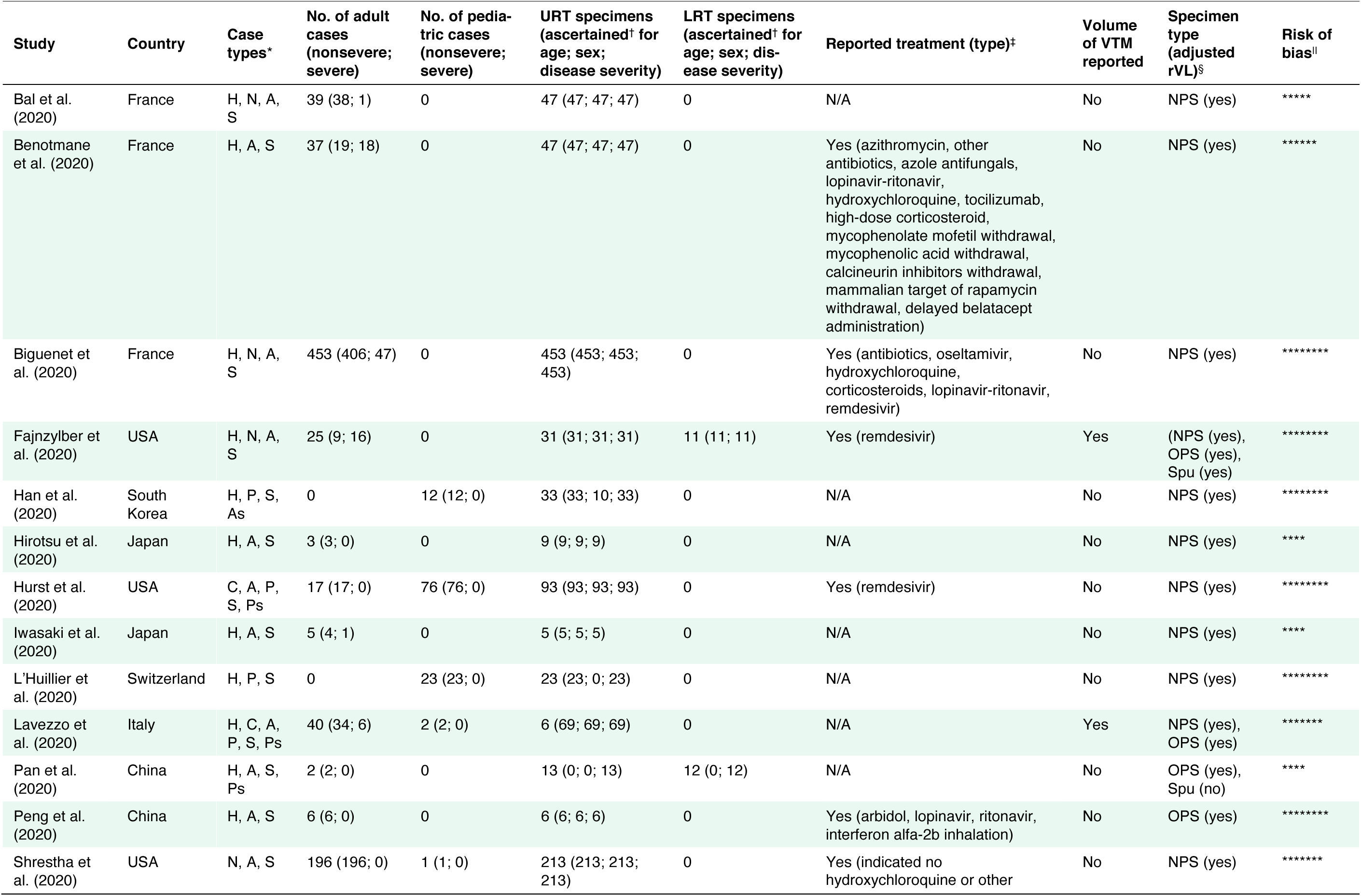
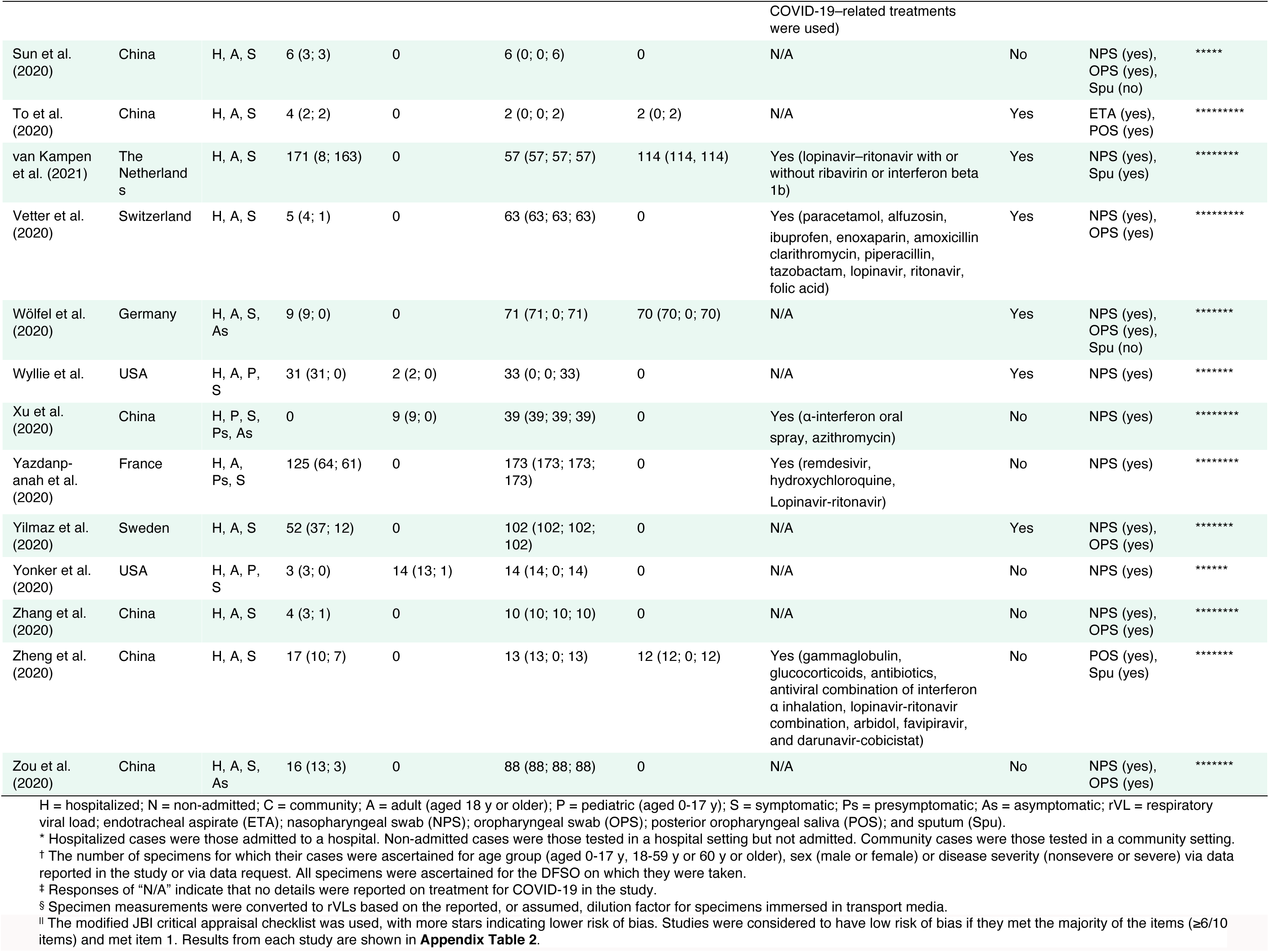
Characteristics of contributing studies.

**Appendix Table 2.**
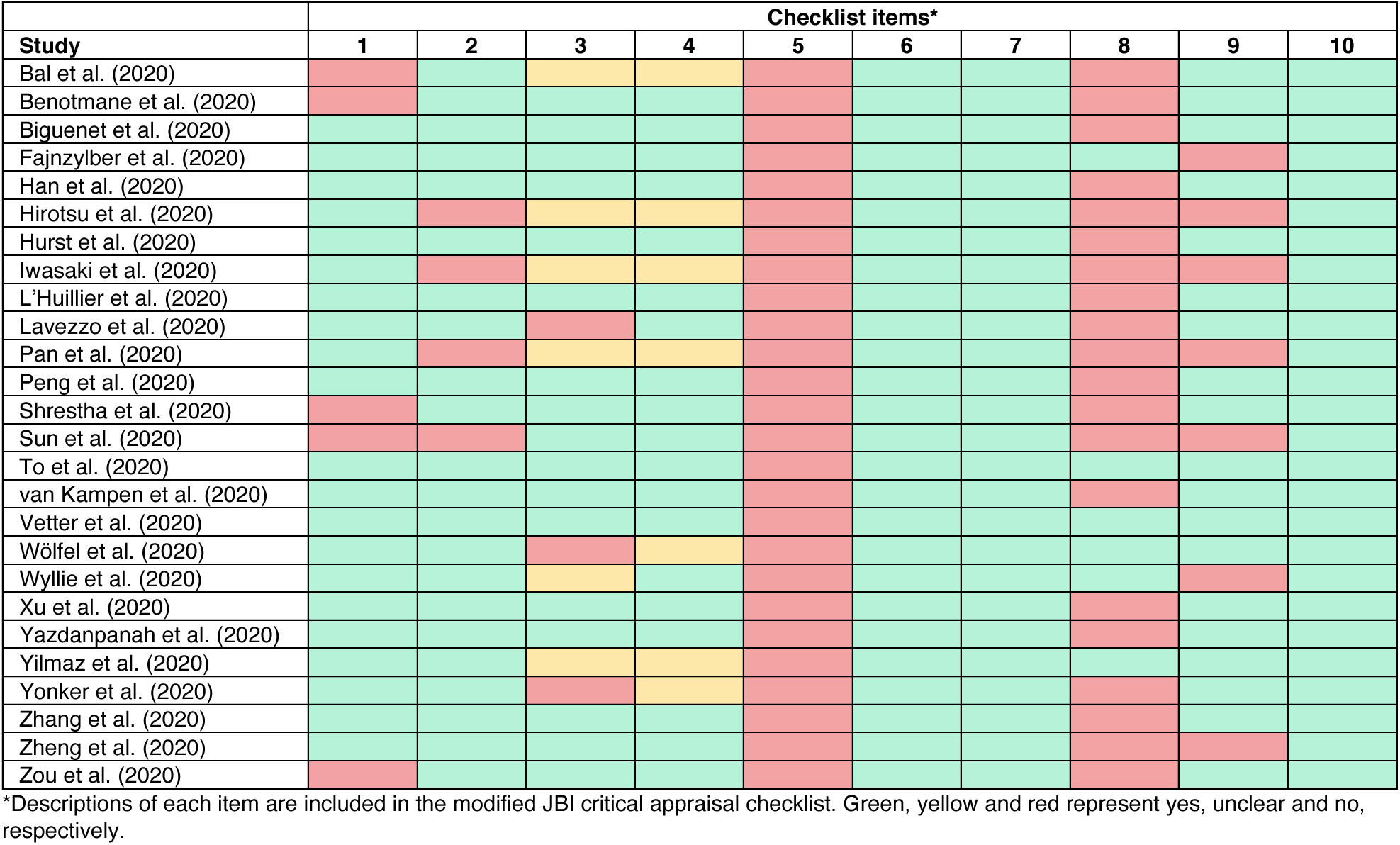
Assessment of risk of bias based on the modified JBI critical appraisal checklist.

**Appendix Table 3.**
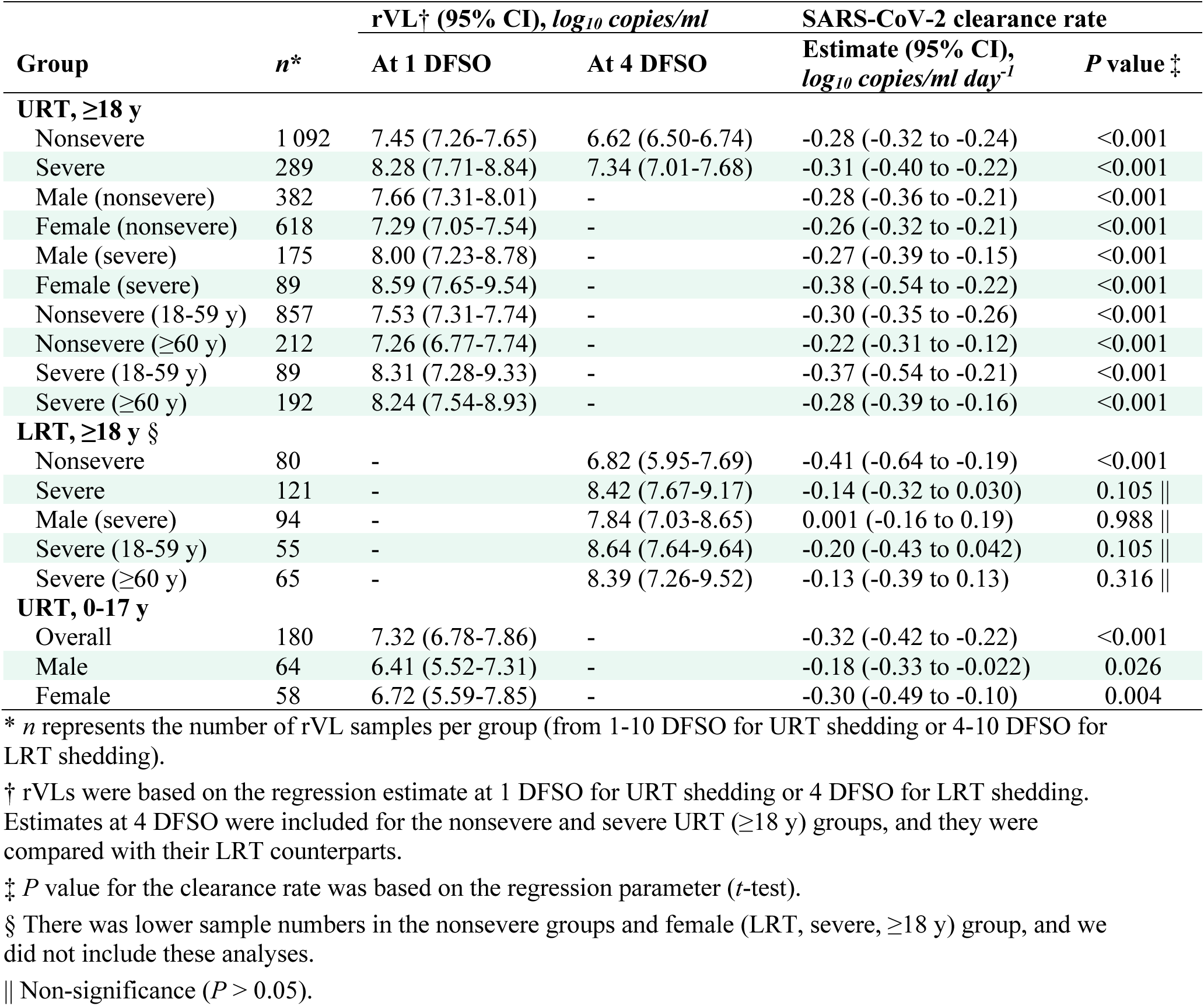
Summary of respiratory shedding levels and dynamics for COVID-19 groups.

**Appendix Table 4.**
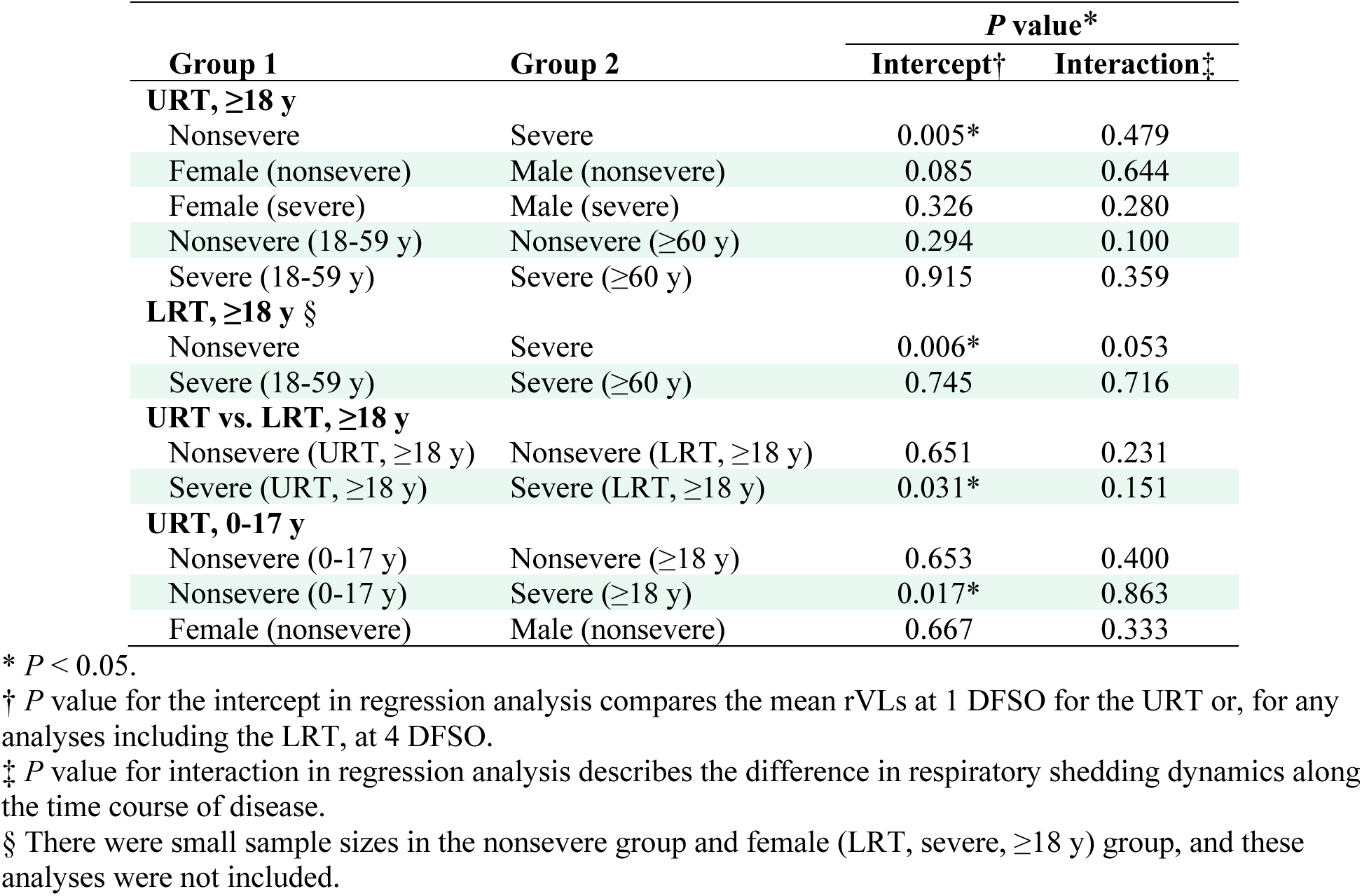
Summary of statistical comparisons on SARS-CoV-2 shedding, across the respiratory tract, COVID-19 severity, sex and age groups

| Group 1 | Group 2 | P value* |  |
| --- | --- | --- | --- |
|  |  | Intercept† | Interaction‡ |
| URT, ≥18 y |  |  |  |
| Nonsevere | Severe | 0.005* | 0.479 |
| Female (nonsevere) | Male (nonsevere) | 0.085 | 0.644 |
| Female (severe) | Male (severe) | 0.326 | 0.280 |
| Nonsevere (18-59 y) | Nonsevere (≥60 y) | 0.294 | 0.100 |
| Severe (18-59 y) | Severe (≥60 y) | 0.915 | 0.359 |
| LRT, ≥18 y § |  |  |  |
| Nonsevere | Severe | 0.006* | 0.053 |
| Severe (18-59 y) | Severe (≥60 y) | 0.745 | 0.716 |
| URT vs. LRT, ≥18 y |  |  |  |
| Nonsevere (URT, ≥18 y) | Nonsevere (LRT, ≥18 y) | 0.651 | 0.231 |
| Severe (URT, ≥18 y) | Severe (LRT, ≥18 y) | 0.031* | 0.151 |
| URT, 0-17 y |  |  |  |
| Nonsevere (0-17 y) | Nonsevere (≥18 y) | 0.653 | 0.400 |
| Nonsevere (0-17 y) | Severe (≥18 y) | 0.017* | 0.863 |
| Female (nonsevere) | Male (nonsevere) | 0.667 | 0.333 |

\* $P < 0.05$ .
† $P$ value for the intercept in regression analysis compares the mean rVLs at 1 DFSO for the URT or, for any analyses including the LRT, at 4 DFSO.
‡ $P$ value for interaction in regression analysis describes the difference in respiratory shedding dynamics along the time course of disease.
§ There were small sample sizes in the nonsevere group and female (LRT, severe, ≥18 y) group, and these analyses were not included.

## Modified JBI critical appraisal checklist

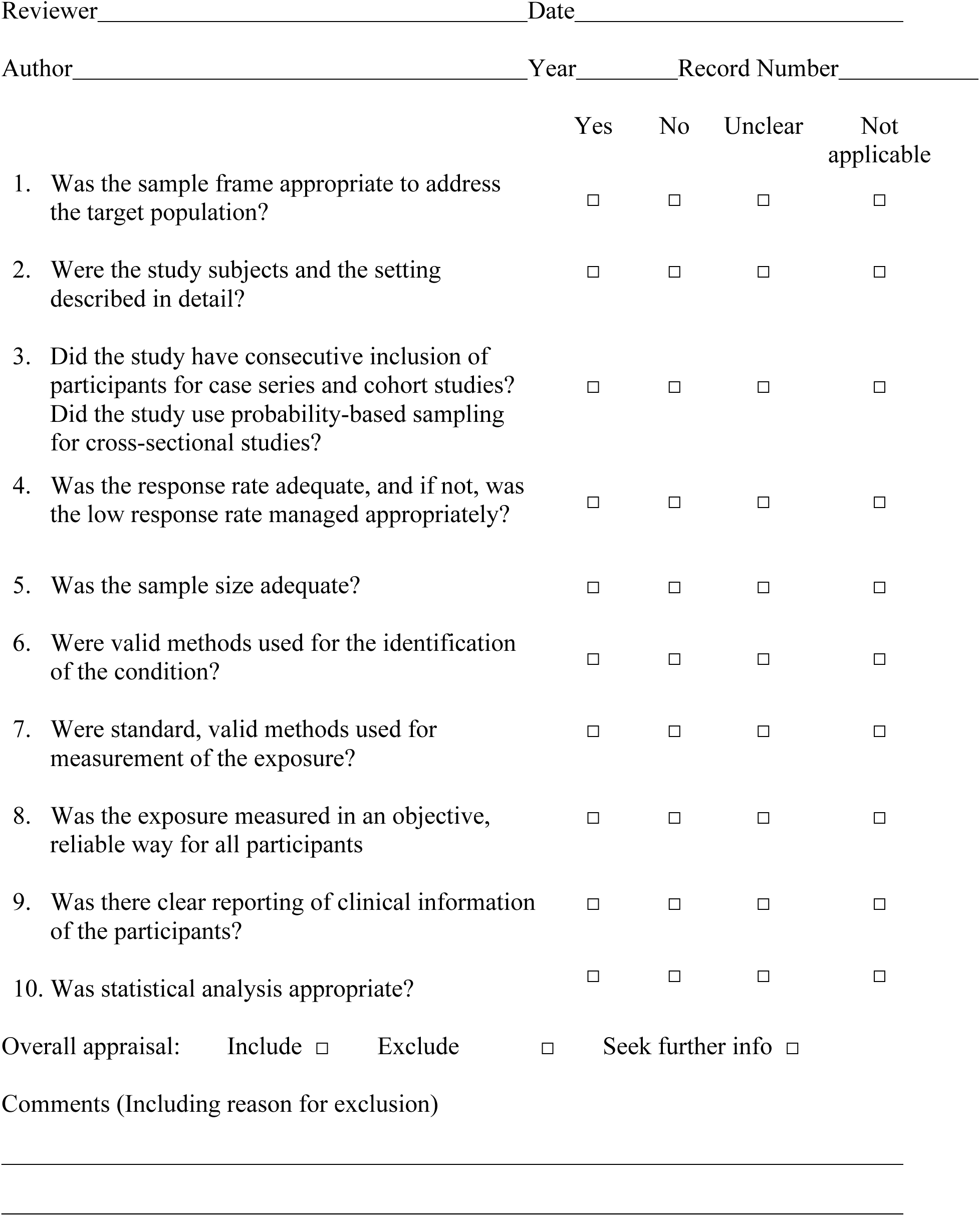

## Tool Guidance

This modified checklist was based on a the JBI Critical Appraisal Checklists for case series, prevalence studies and analytical cross-sectional studies.

1. **Was the sample frame appropriate to address the target population?** This question relies upon knowledge of the broader characteristics of the population of interest and the geographical area. This study broadly investigates the respiratory viral load for the population of interest, which is the general population infected with SARS-CoV-2. The geographical area is not constrained. Sample frames restricted to particular subgroups within the general infected population were considered appropriate if they targeted one of the following groups analysed in our study: asymptomatic, presymptomatic, symptomatic, adult, pediatric, hospitalized, non-admitted, or community.
2. **Were the study subjects and the setting described in detail?** Certain diseases or conditions vary in prevalence across different geographic regions and populations (e.g. Women vs. Men, sociodemographic variables between countries). The study sample should be described in sufficient detail so that other researchers can determine if it is comparable to the population of interest to them
3. **Did the study have consecutive inclusion of participants for case series and cohort studies? Did the study use probability sampling for cross-sectional studies?** Inclusion of consecutive participants for case series and cohort studies yields results at lower risk of bias compared to other sampling methods for these study designs. Use of probability-based sampling methods for cross-sectional studies yields estimates at lower risk of bias compared to other sampling methods for this design. Studies that indicate a consecutive inclusion are more reliable than those that do not. For example, a case series that states ‘we included all patients (24) with osteosarcoma who presented to our clinic between March 2005 and June 2006’ is more reliable than a study that simply states ‘we report a case series of 24 people with osteosarcoma.’
4. **Was the response rate adequate, and if not, was the low response rate managed appropriately?** A large number of dropouts, refusals or “not founds” amongst selected subjects may diminish a study’s validity, as can a low response rates for survey studies. The authors should clearly discuss the response rate and any reasons for non-response and compare persons in the study to those not in the study, particularly with regards to their socio-demographic characteristics. If reasons for non-response appear to be unrelated to the outcome measured and the characteristics of non-responders are comparable to those who do respond in the study, the researchers may be able to justify a more modest response rate.
5. **Was the sample size adequate?** The larger the sample, the narrower will be the confidence interval around the prevalence estimate, making the results more precise. An adequate sample size is important to ensure good precision of the final estimate. The sample size threshold was calculated as follows:

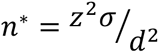

where n* is the sample size threshold, *z* is the z-score for the level of confidence (95%), *σ* is the standard deviation (assumed to be 3 log_10_ copies/ml, a fourth of the full range of rVLs) and *d* is the marginal error (assumed to be 1 log_10_ copies/ml, based on the minimum detection limit for qRT-PCR across studies). This item was met if ≥75% of the included DFSO had ≥46 specimen measurements.
6. **Were valid methods used for the identification of the condition?** Many health problems are not easily diagnosed or defined and some measures may not be capable of including or excluding appropriate levels or stages of the health problem. If the outcomes were assessed based on existing definitions or diagnostic criteria, then the answer to this question is likely to be yes. If the outcomes were assessed using observer reported, or self-reported scales, the risk of over- or under- reporting is increased, and objectivity is compromised. Importantly, determine if the measurement tools used were validated instruments as this has a significant impact on outcome assessment validity.
7. **Were standard, valid methods used for measurement of the exposure?** The study should clearly describe the method of measurement of exposure. Assessing validity requires that a ’gold standard’ is available to which the measure can be compared. The validity of exposure measurement usually relates to whether a current measure is appropriate or whether a measure of past exposure is needed. In this study, standard methods to measure viral load in respiratory specimens are assays quantifying via one of the diagnostic sequences (*Ofr1b*, *N*, *RdRp* and *E* genes) for SARS-CoV-2.
8. **Was the exposure measured in an objective, reliable way for all participants?** The study should clearly describe the procedural aspects of the measurement of exposure as well as factors that can contribute to heterogeneity in measurement. In this study, objective, reliable interpretation of the exposure depends on the use of quantitative calibration; the specification of extraction; determination of the viral load as a standard metric (e.g., copies/ml or equivalent) or in a manner that can be converted to a standard metric; and, if present, specification of the amount of diluent (e.g., viral transport media) used.
9. **Was there clear reporting of clinical information of the participants?** There should be clear reporting of clinical information of the participants such as the following information where relevant: disease status, comorbidities, stage of disease, previous interventions/treatment, results of diagnostic tests, etc. In addition, there should be clear reporting of the number and types (asymptomatic, presymptomatic, symptomatic, adult, pediatric, hospitalized, non-admitted, community, etc.) of cases for measurements within the sampling periods of interest. For studies that include data outside of the infectious period, there should be clear reporting of clinical information for participants for the specimen measurements that were collected from within the infectious period.
10. **Was statistical analysis appropriate?** As with any consideration of statistical analysis, consideration should be given to whether there was a more appropriate alternate statistical method that could have been used. The methods section of studies should be detailed enough for reviewers to identify which analytical techniques were used and whether these were suitable.

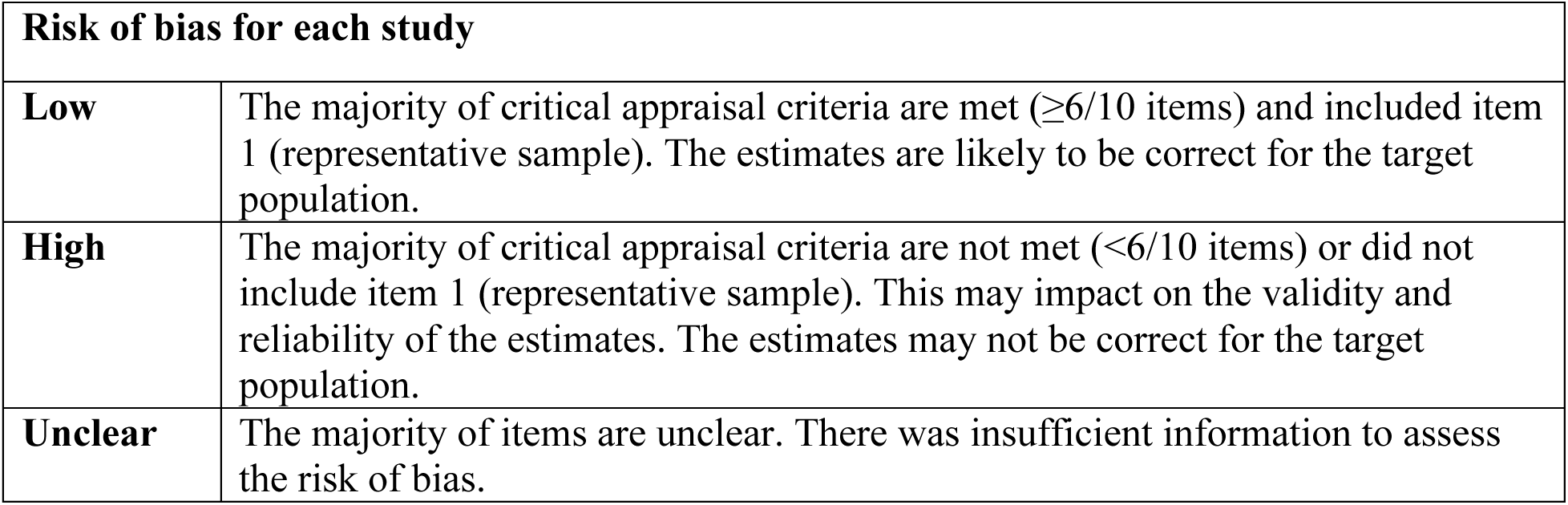

## Notes

### Competing Interest Statement

Dr. Fisman has received honoraria related to work with Pfizer, Astra Zeneca and Seqirus on vaccines for respiratory viruses.

### Funding Statement

This study was supported by NSERC. Mr. Chen was supported by the NSERC Vanier Canada Graduate Scholarship (608544). Dr. Fisman was supported by the Canadian Institutes of Health Research (Canadian COVID-19 Rapid Research Fund, OV4-170360). Dr. Gu was supported by the NSERC Senior Industrial Research Chair.

### Summary of Updates

Updated analyses (and each figure, except for Fig. 1) to show additional information on the comparisons of shedding dynamics of SARS-CoV-2 in the upper and lower respiratory tract

