## Appendix for "SARS-CoV-2 shedding dynamics across the respiratory tract, sex and disease severity for adult and pediatric COVID-19" (from another version of this preprint: version 1, 2021-02-19)

**Appendix Table 1.** Characteristics of contributing studies.

| Study | Country | Case types* | No. of adult cases (nonsevere; severe) | No. of pediatric cases (nonsevere; severe) | URT specimens (ascertained† for age; sex; disease severity) | LRT specimens (ascertained† for age; sex; disease severity) | Reported treatment (type)‡ | Volume of VTM reported | Specimen type (adjusted rVL)§ | Risk of bias‖ |
|---|---|---|---|---|---|---|---|---|---|---|
| Bal et al. (2020) | France | H, N, A, S | 39 (38; 1) | 0 | 47 (47; 47; 47) | 0 | N/A | No | NPS (yes) | ***** |
| Benotmane et al. (2020) | France | H, A, S | 37 (19; 18) | 0 | 47 (47; 47; 47) | 0 | Yes (azithromycin, other antibiotics, azole antifungals, lopinavir-ritonavir, hydroxychloroquine, tocilizumab, high-dose corticosteroid, mycophenolate mofetil withdrawal, mycophenolic acid withdrawal, calcineurin inhibitors withdrawal, mammalian target of rapamycin withdrawal, delayed belatacept administration) | No | NPS (yes) | ****** |
| Biguenet et al. (2020) | France | H, N, A, S | 453 (406; 47) | 0 | 453 (453; 453; 453) | 0 | Yes (antibiotics, oseltamivir, hydroxychloroquine, corticosteroids, lopinavir-ritonavir, remdesivir) | No | NPS (yes) | ******** |
| Fajnzylber et al. (2020) | USA | H, N, A, S | 25 (9; 16) | 0 | 31 (31; 31; 31) | 11 (11; 11) | Yes (remdesivir) | Yes | (NPS (yes), OPS (yes), Spu (yes) | ******** |
| Han et al. (2020) | South Korea | H, P, S, As | 0 | 12 (12; 0) | 33 (33; 10; 33) | 0 | N/A | No | NPS (yes) | ******** |
| Hirotsu et al. (2020) | Japan | H, A, S | 3 (3; 0) | 0 | 9 (9; 9; 9) | 0 | N/A | No | NPS (yes) | **** |
| Hurst et al. (2020) | USA | C, A, P, S, Ps | 17 (17; 0) | 76 (76; 0) | 93 (93; 93; 93) | 0 | Yes (remdesivir) | No | NPS (yes) | ******** |
| Iwasaki et al. (2020) | Japan | H, A, S | 5 (4; 1) | 0 | 5 (5; 5; 5) | 0 | N/A | No | NPS (yes) | **** |
| L'Huillier et al. (2020) | Switzerland | H, P, S | 0 | 23 (23; 0) | 23 (23; 0; 23) | 0 | N/A | No | NPS (yes) | ******** |
| Lavezzo et al. (2020) | Italy | H, C, A, P, S, Ps | 40 (34; 6) | 2 (2; 0) | 6 (69; 69; 69) | 0 | N/A | Yes | NPS (yes), OPS (yes) | ******* |
| Pan et al. (2020) | China | H, A, S, Ps | 2 (2; 0) | 0 | 13 (0; 0; 13) | 12 (0; 12) | N/A | No | OPS (yes), Spu (no) | **** |
| Peng et al. (2020) | China | H, A, S | 6 (6; 0) | 0 | 6 (6; 6; 6) | 0 | Yes (arbidol, lopinavir, ritonavir, interferon alfa-2b inhalation) | No | OPS (yes) | ******** |
| Shrestha et al. (2020) | USA | N, A, S | 196 (196; 0) | 1 (1; 0) | 213 (213; 213; 213) | 0 | Yes (indicated no hydroxychloroquine or other COVID-19–related treatments were used) | No | NPS (yes) | ******* |

| | | | | | | | | | | |
|---|---|---|---|---|---|---|---|---|---|---|
| Sun et al. (2020) | China | H, A, S | 6 (3; 3) | 0 | 6 (0; 0; 6) | 0 | N/A | No | NPS (yes), OPS (yes), Spu (no) | ***** |
| To et al. (2020) | China | H, A, S | 4 (2; 2) | 0 | 2 (0; 0; 2) | 2 (0; 2) | N/A | Yes | ETA (yes), POS (yes) | ********* |
| van Kampen et al. (2021) | The Netherland s | H, A, S | 171 (8; 163) | 0 | 57 (57; 57; 57) | 114 (114, 114) | Yes (lopinavir–ritonavir with or without ribavirin or interferon beta 1b) | Yes | NPS (yes), Spu (yes) | ******** |
| Vetter et al. (2020) | Switzerland | H, A, S | 5 (4; 1) | 0 | 63 (63; 63; 63) | 0 | Yes (paracetamol, alfuzosin, ibuprofen, enoxaparin, amoxicillin clarithromycin, piperacillin, tazobactam, lopinavir, ritonavir, folic acid) | Yes | NPS (yes), OPS (yes) | ********* |
| Wölfel et al. (2020) | Germany | H, A, S, As | 9 (9; 0) | 0 | 71 (71; 0; 71) | 70 (70; 0; 70) | N/A | Yes | NPS (yes), OPS (yes), Spu (no) | ******* |
| Wyllie et al. | USA | H, A, P, S | 31 (31; 0) | 2 (2; 0) | 33 (0; 0; 33) | 0 | N/A | Yes | NPS (yes) | ******* |
| Xu et al. (2020) | China | H, P, S, Ps, As | 0 | 9 (9; 0) | 39 (39; 39; 39) | 0 | Yes (α-interferon oral spray, azithromycin) | No | NPS (yes) | ******** |
| Yazdanp-anah et al. (2020) | France | H, A, Ps, S | 125 (64; 61) | 0 | 173 (173; 173; 173) | 0 | Yes (remdesivir, hydroxychloroquine, Lopinavir-ritonavir) | No | NPS (yes) | ******** |
| Yilmaz et al. (2020) | Sweden | H, A, S | 52 (37; 12) | 0 | 102 (102; 102; 102) | 0 | N/A | Yes | NPS (yes), OPS (yes) | ******* |
| Yonker et al. (2020) | USA | H, A, P, S | 3 (3; 0) | 14 (13; 1) | 14 (14; 0; 14) | 0 | N/A | No | NPS (yes) | ****** |
| Zhang et al. (2020) | China | H, A, S | 4 (3; 1) | 0 | 10 (10; 10; 10) | 0 | N/A | No | NPS (yes), OPS (yes) | ******** |
| Zheng et al. (2020) | China | H, A, S | 17 (10; 7) | 0 | 13 (13; 0; 13) | 12 (12; 0; 12) | Yes (gammaglobulin, glucocorticoids, antibiotics, antiviral combination of interferon α inhalation, lopinavir-ritonavir combination, arbidol, favipiravir, and darunavir-cobicistat) | No | POS (yes), Spu (yes) | ******* |
| Zou et al. (2020) | China | H, A, S, As | 16 (13; 3) | 0 | 88 (88; 88; 88) | 0 | N/A | No | NPS (yes), OPS (yes) | ******* |

H = hospitalized; N = non-admitted; C = community; A = adult (aged 18 y or older); P = pediatric (aged 0-17 y); S = symptomatic; Ps = presymptomatic; As = asymptomatic; rVL = respiratory viral load; endotracheal aspirate (ETA); nasopharyngeal swab (NPS); oropharyngeal swab (OPS); posterior oropharyngeal saliva (POS); and sputum (Spu).

* Hospitalized cases were those admitted to a hospital. Non-admitted cases were those tested in a hospital setting but not admitted. Community cases were those tested in a community setting.

† The number of specimens for which their cases were ascertained for age cohort (aged 0-17 y, 18-59 y or 60 y or older), sex (male or female) or disease severity (nonsevere or severe) via data reported in the study or via data request. All specimens were ascertained for the DFSO on which they were taken.

‡ Responses of "N/A" indicate that no details were reported on treatment for COVID-19 in the study.

§ Specimen measurements were converted to rVLs based on the reported, or assumed, dilution factor for specimens immersed in transport media.

‖ The modified JBI critical appraisal checklist was used, with more stars indicating lower risk of bias. Studies were considered to have low risk of bias if they met the majority of the items (≥6/10 items) and met item 1. Results from each study are shown in **Appendix Table 2**.

**Appendix Table 2.** Assessment of risk of bias based on the modified JBI critical appraisal checklist.

| | Checklist items* | | | | | | | | | |
|---|---|---|---|---|---|---|---|---|---|---|
| **Study** | **1** | **2** | **3** | **4** | **5** | **6** | **7** | **8** | **9** | **10** |
| Bal et al. (2020) | Red | Green | Yellow | Yellow | Red | Green | Green | Red | Green | Green |
| Benotmane et al. (2020) | Red | Green | Green | Green | Red | Green | Green | Red | Green | Green |
| Biguenet et al. (2020) | Green | Green | Green | Green | Red | Green | Green | Red | Green | Green |
| Fajnzylber et al. (2020) | Green | Green | Green | Green | Red | Green | Green | Green | Red | Green |
| Han et al. (2020) | Green | Green | Green | Green | Red | Green | Green | Red | Green | Green |
| Hirotsu et al. (2020) | Green | Red | Yellow | Yellow | Red | Green | Green | Red | Red | Green |
| Hurst et al. (2020) | Green | Green | Green | Green | Red | Green | Green | Red | Green | Green |
| Iwasaki et al. (2020) | Green | Red | Yellow | Yellow | Red | Green | Green | Red | Red | Green |
| L'Huillier et al. (2020) | Green | Green | Green | Green | Red | Green | Green | Red | Green | Green |
| Lavezzo et al. (2020) | Green | Green | Red | Green | Red | Green | Green | Red | Green | Green |
| Pan et al. (2020) | Green | Red | Yellow | Yellow | Red | Green | Green | Red | Red | Green |
| Peng et al. (2020) | Green | Green | Green | Green | Red | Green | Green | Red | Green | Green |
| Shrestha et al. (2020) | Red | Green | Green | Green | Red | Green | Green | Red | Green | Green |
| Sun et al. (2020) | Red | Red | Green | Green | Red | Green | Green | Red | Red | Green |
| To et al. (2020) | Green | Green | Green | Green | Red | Green | Green | Green | Green | Green |
| van Kampen et al. (2020) | Green | Green | Green | Green | Red | Green | Green | Red | Green | Green |
| Vetter et al. (2020) | Green | Green | Green | Green | Red | Green | Green | Green | Green | Green |
| Wölfel et al. (2020) | Green | Green | Red | Yellow | Red | Green | Green | Green | Green | Green |
| Wyllie et al. (2020) | Green | Green | Yellow | Green | Red | Green | Green | Green | Red | Green |
| Xu et al. (2020) | Green | Green | Green | Green | Red | Green | Green | Red | Green | Green |
| Yazdanpanah et al. (2020) | Green | Green | Green | Green | Red | Green | Green | Red | Green | Green |
| Yilmaz et al. (2020) | Green | Green | Yellow | Yellow | Red | Green | Green | Green | Green | Green |
| Yonker et al. (2020) | Green | Green | Red | Yellow | Red | Green | Green | Red | Green | Green |
| Zhang et al. (2020) | Green | Green | Green | Green | Red | Green | Green | Red | Green | Green |
| Zheng et al. (2020) | Green | Green | Green | Green | Red | Green | Green | Red | Red | Green |
| Zou et al. (2020) | Red | Green | Green | Green | Red | Green | Green | Red | Green | Green |

*Descriptions of each item are included in the modified JBI critical appraisal checklist (**Supplement Table 6**). Green, yellow and red represent yes, unclear and no, respectively.

**Appendix Table 3.** Summary of respiratory shedding levels and dynamics for COVID-19 cohorts.

| | | rVL† (95% CI), *$log_{10}$ copies/ml* | | SARS-CoV-2 clearance rate | |
|---|---|---|---|---|---|
| **Cohort*** | ***n*** | **At 1 DFSO** | **At 4 DFSO** | **Estimate (95% CI),** ***$log_{10}$ copies/ml $day^{-1}$*** | ***P* value ‡** |
| **URT, ≥18 y** | | | | | |
| Nonsevere | 1 092 | 7.45 (7.26-7.65) | 6.62 (6.50-6.74) | -0.28 (-0.32 to -0.24) | <0.001 |
| Severe | 289 | 8.28 (7.71-8.84) | 7.34 (7.01-7.68) | -0.31 (-0.40 to -0.22) | <0.001 |
| Male (nonsevere) | 382 | 7.66 (7.31-8.01) | - | -0.28 (-0.36 to -0.21) | <0.001 |
| Female (nonsevere) | 618 | 7.29 (7.05-7.54) | - | -0.26 (-0.32 to -0.21) | <0.001 |
| Male (severe) | 175 | 8.00 (7.23-8.78) | - | -0.27 (-0.39 to -0.15) | <0.001 |
| Female (severe) | 89 | 8.59 (7.65-9.54) | - | -0.38 (-0.54 to -0.22) | <0.001 |
| Nonsevere (18-59 y) | 857 | 7.53 (7.31-7.74) | - | -0.30 (-0.35 to -0.26) | <0.001 |
| Nonsevere (≥60 y) | 212 | 7.26 (6.77-7.74) | - | -0.22 (-0.31 to -0.12) | <0.001 |
| Severe (18-59 y) | 89 | 8.31 (7.28-9.33) | - | -0.37 (-0.54 to -0.21) | <0.001 |
| Severe (≥60 y) | 192 | 8.24 (7.54-8.93) | - | -0.28 (-0.39 to -0.16) | <0.001 |
| **LRT, ≥18 y** § | | | | | |
| Nonsevere | 80 | - | 6.82 (5.95-7.69) | -0.41 (-0.64 to -0.19) | <0.001 |
| Severe | 121 | - | 8.42 (7.67-9.17) | -0.14 (-0.32 to 0.030) | 0.105 ‖ |
| Male (severe) | 94 | - | 7.84 (7.03-8.65) | 0.001 (-0.16 to 0.19) | 0.988 ‖ |
| Severe (18-59 y) | 55 | - | 8.64 (7.64-9.64) | -0.20 (-0.43 to 0.042) | 0.105 ‖ |
| Severe (≥60 y) | 65 | - | 8.39 (7.26-9.52) | -0.13 (-0.39 to 0.13) | 0.316 ‖ |
| **URT, 0-17 y** | | | | | |
| Overall | 180 | 7.32 (6.78-7.86) | - | -0.32 (-0.42 to -0.22) | <0.001 |
| Male | 64 | 6.41 (5.52-7.31) | - | -0.18 (-0.33 to -0.022) | 0.026 |
| Female | 58 | 6.72 (5.59-7.85) | - | -0.30 (-0.49 to -0.10) | 0.004 |

DFSO = days from symptom onset; LRT = lower respiratory tract; nonsevere = asymptomatic, mild and moderate COVID-19; rVL = respiratory viral load; severe = severe and critical COVID-19; URT = upper respiratory tract.

* Each regression analysis was shown in **Figures 2 to 4**. *n* represents the number of rVL samples per cohort (from 1-10 DFSO for URT shedding or 4-10 DFSO for LRT shedding).

† rVLs were based on the regression estimate at 1 DFSO for URT shedding or 4 DFSO for LRT shedding. Estimates at 4 DFSO were included for the nonsevere and severe URT (≥18 y) cohorts, and they were compared with their LRT counterparts.

‡ *P* value for the clearance rate was based on the regression parameter (*t*-test).

§ There was lower sample numbers in the nonsevere cohorts and female (LRT, severe, ≥18 y) cohort, and we did not include these analyses.

‖ Non-significance ($P > 0.05$).

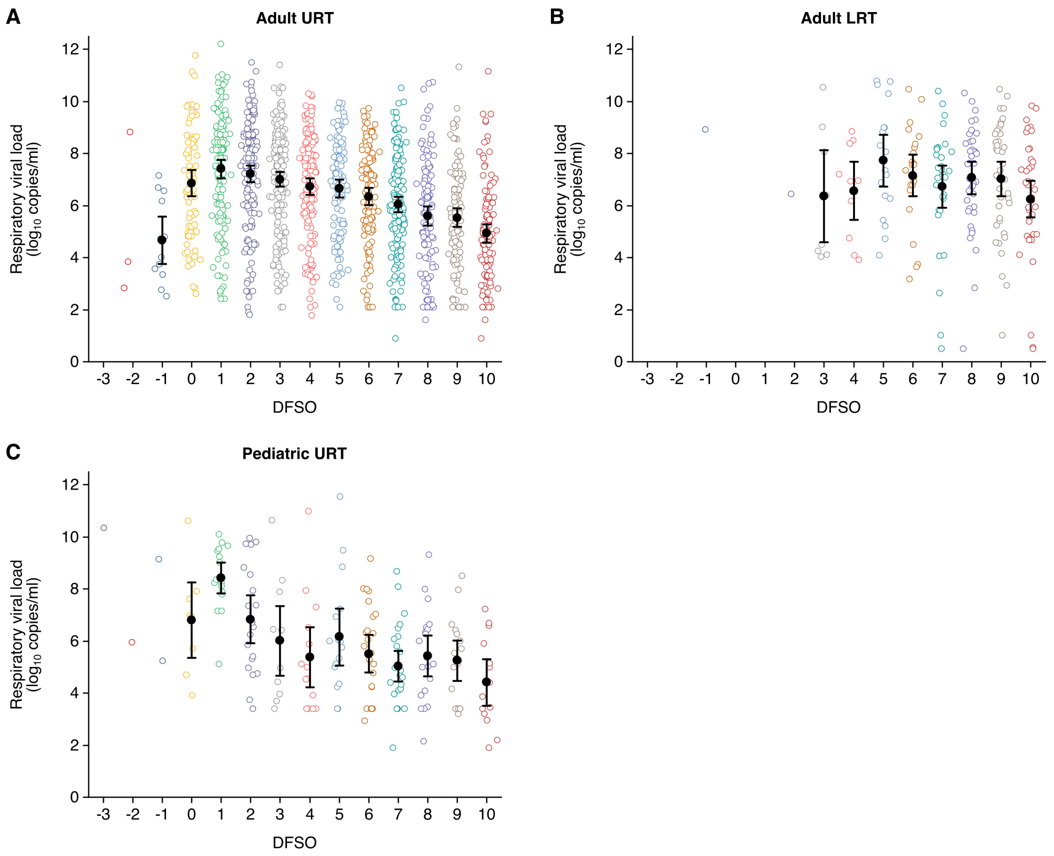


**Appendix Figure. Summary of rVLs in the systematic dataset. A**. URT rVLs for adult COVID-19 cases (from -3 to 10 days from symptom onset [DFSO], *n* = 0, 3, 12, 71, 145, 159, 176, 152, 114, 123, 161, 125, 110, and 117 samples per DFSO). **B**. LRT rVLs for adult COVID-19 cases (from -3 to 10 DFSO, *n* = 0, 0, 1, 0, 0, 1, 8, 10, 18, 23, 31, 39, 40, and 40 samples per DFSO). **C**. URT rVLs for pediatric COVID-19 cases (from -3 to 10 DFSO, *n* = 1, 1, 2, 8, 17, 20, 11, 14, 16, 25, 26, 20, 17, and 14 samples per DFSO). Data were collected within the estimated infectious period of SARS-CoV-2 (-3 to 10 DFSO). The systematic search found no quantitative specimen measurements from the LRT for pediatric COVID-19. Open circles show rVL data. Filled circles and bars depict mean estimates and 95% CIs, respectively.
