## Supplement for "SARS-CoV-2 shedding dynamics across the respiratory tract, sex and disease severity for adult and pediatric COVID-19" (from another version of this preprint: version 1, 2021-02-19)

**Additional Details on Data Searches**

As it was based on a prior study, the protocol searched for respiratory specimen measurements from three diseases (COVID-19, severe acute respiratory syndrome [SARS] and influenza A/H1N1pdm09) during the estimated infectious period of each virus. Hence, our search was inclusive of the desired data for SARS-CoV-2 specimen measurements for COVID-19 cases, and we included only these data.

The search strategy, which included subject headings and keywords, was developed by a librarian (Z.P.) and had 3 main concepts (disease, specimen type and outcome); each concept was combined using the appropriate Boolean operators. The search was tested against a sample set of known articles that were pre-identified. We searched, without the use of filters or language restrictions, the following sources: MEDLINE (via Ovid, 1946 to 20 November 2020); EMBASE (via Ovid, 1974 to 20 November, 2020); Cochrane Central Register of Controlled Trials (via Ovid, 1991 to 20 November, 2020); Web of Science Core Collection (which includes: Science Citation Index Expanded, 1900 to 20 November 2020; Social Sciences Citation Index, 1900 to 20 November 2020; Arts & Humanities Citation Index, 1975 to 20 November 2020; Conference Proceedings Citation Index - Science, 1990 to 20 November 2020; Conference Proceedings Citation Index - Social Sciences & Humanities, 1990 to 20 November 2020; Emerging Sources Citation Index, 2015 to 20 November 2020); and medRxiv and bioRxiv (both searched through Google Scholar via the Publish or Perish program).

The line-by-line search strategies for all databases are included in **Supplement Tables 1** to **5**. The search results were exported from each database and uploaded to the Covidence online system for deduplication and screening.

**Additional Details on Study Selection**

The search included papers that (i) reported positive, quantitative measurements (copies/ml or an equivalent metric) of SARS-CoV-2 in human respiratory specimens (endotracheal aspirate, nasopharyngeal aspirate, nasopharyngeal swabs, oropharyngeal swabs, posterior oropharyngeal saliva and sputum) from COVID-19, cases; (ii) reported data that could be extracted from the estimated infectious period of SARS-CoV-2 (defined as -3 to +10 DFSO for symptomatic cases and 0 to +10 days from the day of laboratory diagnosis for asymptomatic cases); and (iii) reported data for two or more cases with laboratory-confirmed COVID-19 based on World Health Organization case definitions. Quantitative specimen measurements were considered after RNA extraction for diagnostic sequences of SARS-CoV-2 (*Ofr1b*, *N*, *RdRp* and *E* genes).

Studies were excluded, in the following order, if they (i) studied an ineligible disease; (ii) had an ineligible study design, including those that were reviews of evidence (e.g., scoping, systematic or narrative), did not include primary clinical human data, reported data for less than two cases due to an increased risk of selection bias, were incomplete (e.g., ongoing clinical trials), did not report an RNA extraction step before measurement or were studies of environmental samples; (iii) reported an ineligible metric for specimen concentration (e.g., qualitative RT-PCR or cycle threshold values without calibration included in the study, or quantitative measurements for which the DFSO of collection was unknown); (iv) reported quantitative measurements from an ineligible specimen type (e.g., blood specimens, pooled specimens or self-collected posterior oropharyngeal saliva or sputum specimens in the absence of a healthcare professional); (v) reported an ineligible sampling period (consisted entirely of data that could not be extracted from within the infectious period); or (vi) were duplicates of an included study (e.g., preprinted version of a published paper or duplicates not identified by

Covidence). When the above criteria were met, we included data from control groups receiving standard of care in interventional studies but excluded data from the intervention group. Patients in the intervention group are, by definition, systematically different from general case populations because they receive therapies not being widely used for treatment, which may influence virus concentrations. Interventional studies examining the comparative effectiveness of two or more treatments were excluded for the same reason. For preprinted studies that were published as journal articles before the submission date of this manuscript, we included the citation for the journal article.

**Development of the Modified JBI Critical Appraisal Checklist**

The modified critical appraisal checklist used in this study (**Supplement Table 6**) was based on a modified of the Joanna Briggs Institute (JBI) tools for case series, analytical cross-sectional studies and prevalence studies (1-3). To evaluate the sample size in a study, we used the following calculation:

$$n^* = \frac{z^2\sigma}{d^2}, \qquad (1)$$

where $n^*$ is the sample size threshold, $z$ is the z-score for the level of confidence (95%), $\sigma$ is the standard deviation (assumed to be 3 $\log_{10}$ copies/ml, one quarter of the full range of rVLs) and $d$ is the marginal error (assumed to be 1 $\log_{10}$ copies/ml, based on the minimum detection limit for qRT-PCR across studies) (4). The modified JBI critical appraisal checklist is shown in **Supplement Table 6**. Items were judged with responses to data inquiries, if authors responded.

**Supplement Table 1.** Search strategy used for MEDLINE.

| Database: **Ovid MEDLINE® and Epub Ahead of Print, In-Process & Other Non-Indexed Citations and Daily** 1946 to Nov 20, 2020 | |
|---|---|
| **#** | **Searches** |
| 1 | exp Coronavirus/ |
| 2 | exp Coronavirus Infections/ |
| 3 | exp Betacoronavirus/ |
| 4 | (coronavirus* or corona virus* or betacoronavirus* or OC43 or NL63 or 229E or HKU1 or HcoV* or ncov* or covid* or sars-cov* or sarscov* or Sars-coronavirus* or Severe Acute Respiratory Syndrome* or sudden acute respiratory syndrome*).tw,kf. |
| 5 | (2019nCov* or 2019-novel CoV or corona or covid19 or ((novel or new or nouveau) adj2 (CoV or Pandemi*))).tw,kf. |
| 6 | (pneumonia.tw,kf. Or exp pneumonia/) and (Wuhan or Hubei).tw,kf. |
| 7 | COVID-19.rx,px,ox. Or severe acute respiratory syndrome coronavirus 2.os. |
| 8 | exp Influenza A Virus, H1N1 Subtype/ |
| 9 | ("A/H1N1*" or H1N1* or pdm09 or ((influenza or virus or pandemic) adj4 "2009") or influenza A or swine flu).tw,kf. |
| 10 | 1 or 2 or 3 or 4 or 5 or 6 or 7 or 8 or 9 |
| 11 | ((respiratory adj3 (specimen* or sample* or swab*)) or sputum or nares or endotrachea* or endotrache* or endotra* or ((nasal or oral* or throat) adj3 (swab* or sample* or smear* or specimen*)) or NPS or OPS or ((endotrachea* or endotracheal*) adj2 aspirat*) or NPA or ETA or (deep adj4 saliva) or POS or "swab sample*" or "flocked swab*").tw,kf. |
| 12 | Nasal cavity/vi |
| 13 | Sputum/vi |
| 14 | Nasopharynx/vi |
| 15 | Oropharynx/vi |
| 16 | *Saliva/vi |
| 17 | Pharynx/vi |
| 18 | (clinical adj2 (sample* or specimen*)).tw,kf. |
| 19 | ("RT-PCR" or "RTPCR" or "ddPCR" or "polymerase chain reaction").tw,kf. |
| 20 | Influenza, Human/vi |
| 21 | exp Coronavirus Infections/vi or exp Coronavirus/vi or exp Betacoronavirus/vi |
| 22 | polymerase chain reaction/ or multiplex polymerase chain reaction/ or real-time polymerase chain reaction/ or reverse transcriptase polymerase chain reaction/ |
| 23 | 11 or 12 or 13 or 14 or 15 or 16 or 17 or 18 or 19 or 20 or 21 or 22 |
| 24 | 10 and 23 |
| 25 | (vir* load* or vir* shed* or vir* burden or vir* titer* or vir* titre* or (vir* adj2 count*)).tw,kf. |
| 26 | (((copies or copy) adj2 (ml* or milli* or microl*)) or ((RNA* or vir*) adj2 concentration*)).tw,kf. |
| 27 | ((calibration adj1 curve*) or (standard adj1 curve*)).tw,kf. |

| | |
|---|---|
| 28 | ((ct* adj1 value*) or cycle threshold or (copies adj2 test*) or (copy adj2 test*) or ((copy or copies) adj2 number*)).tw,kf. |
| 29 | Viral load/ |
| 30 | Viral shedding/ |
| 31 | (“copy/m*” or “copies/m*” or “copy/test*” or “copies/test*”).tw,kf. |
| 32 | ((test or diagnos*) adj2 sensitiv*).tw,kf. |
| 33 | 25 or 26 or 27 or 28 or 29 or 30 or 31 or 32 |
| 34 | 24 and 33 |
| 35 | animals/ not humans/ |
| 36 | 34 not 35 |

**Supplement Table 2.** Search strategy used for EMBASE.

| Database: **Embase** 1974 to Nov 20, 2020 | |
|---|---|
| **#** | **Searches** |
| 1 | coronavirus infection/ or severe acute respiratory syndrome/ |
| 2 | COVID 19/ |
| 3 | coronavirus disease 2019/ |
| 4 | severe acute respiratory syndrome coronavirus 2/ or SARS coronavirus 2/ |
| 5 | SARS coronavirus/ or betacoronavirus/ |
| 6 | "influenza a virus (h1n1)"/ |
| 7 | 2009 h1n1 influenza/ or "influenza a (h1n1)"/ |
| 8 | (coronavirus* or corona virus* or betacoronavirus* or OC43 or NL63 or 229E or HKU1 or HcoV* or ncov* or covid* or sars-cov* or sarscov* or Sars-coronavirus* or Severe Acute Respiratory Syndrome* or Sudden acute respiratory syndrome*).tw,kw. |
| 9 | (2019nCov* or 2019-novel CoV or corona or covid19 or ((novel or new or nouveau) adj2 (CoV or Pandemi*))).tw,kw. |
| 10 | (pneumonia.tw,kw. Or exp pneumonia/) and (Wuhan or Hubei).tw,kw. |
| 11 | (covid or SARS or H1N1 or coronavirus).ox. |
| 12 | ("A/H1N1*" or H1N1* or pdm09 or ((influenza or virus or pandemic) adj4 "2009") or influenza A or swine flu).tw,kw. |
| 13 | 1 or 2 or 3 or 4 or 5 or 6 or 7 or 8 or 9 or 10 or 11 or 12 |
| 14 | nose smear/ |
| 15 | smear/ or nose smear/ or sputum smear/ |
| 16 | *sputum/ |
| 17 | *nasopharynx/ or *pharynx/ |
| 18 | throat culture/ |
| 19 | saliva analysis/ |
| 20 | *oropharynx/ |
| 21 | exp nasopharyngeal aspiration/ |
| 22 | real time polymerase chain reaction/ or real time reverse transcription polymerase chain reaction/ or reverse transcription polymerase chain reaction/ |
| 23 | (clinical adj2 (sample* or specimen*)).tw,kw. |
| 24 | ((respiratory adj3 (specimen* or sample* or swab*)) or sputum or nares or endotrachea* or endotrache* or endotra* or ((nasal or oral* or throat) adj3 (swab* or sample* or smear* or specimen*)) or NPS or OPS or ((endotrachea* or endotracheal*) adj2 aspirat*) or NPA or ETA or (deep adj4 saliva) or POS or "swab sample*" or "flocked swab*").tw,kw. |
| 25 | ("RT-PCR" or "RTPCR" or "ddPCR" or "polymerase chain reaction").tw,kw. |
| 26 | *2009 H1N1 influenza/di or coronavirus disease 2019/di or Coronavirus infection/di |

| | |
|---|---|
| 27 | 14 or 15 or 16 or 17 or 18 or 19 or 20 or 21 or 22 or 23 or 24 or 25 or 26 |
| 28 | 13 and 27 |
| 29 | virus load/ |
| 30 | virus shedding/ |
| 31 | (vir* load* or vir* shed* or vir* burden or vir* titer* or vir* titre* or (vir* adj2 count*)).tw,kw. |
| 32 | (((copies or copy) adj2 (ml* or milli* or microl*)) or ((RNA* or vir*) adj2 concentration*)).tw,kw. |
| 33 | ((calibration adj1 curve*) or (standard adj1 curve*)).tw,kw. |
| 34 | ((ct* adj1 value*) or cycle threshold or (copies adj2 test*) or (copy adj2 test*) or ((copy or copies) adj2 number*)).tw,kw. |
| 35 | ("copy/m*" or "copies/m*" or "copy/test*" or "copies/test*").tw,kw. |
| 36 | ((test or diagnos*) adj2 sensitiv*).tw,kw. |
| 37 | 29 or 30 or 31 or 32 or 33 or 34 or 35 or 36 |
| 38 | 28 and 37 |
| 39 | exp animal/ not human/ |
| 40 | 38 not 39 |

**Supplement Table 3.** Search strategy used for Cochrane Central.

| Database(s): **EBM Reviews – Cochrane Central Register of Controlled Trials** Nov 20, 2020 | |
|---|---|
| **#** | **Searches** |
| 1 | exp Coronavirus/ |
| 2 | exp Coronavirus Infections/ |
| 3 | betacoronavirus/ |
| 4 | (coronavirus* or corona virus* or betacoronavirus* or OC43 or NL63 or 229E or HKU1 or HcoV* or ncov* or covid* or sars-cov* or sarscov* or Sars-coronavirus* or Severe Acute Respiratory Syndrome* or sudden acute respiratory syndrome*).tw,kw. |
| 5 | (2019nCov* or 2019-novel CoV or corona or covid19 or ((novel or new or nouveau) adj2 (CoV or Pandemi*))).tw,kw. |
| 6 | (pneumonia.tw,kw. Or exp pneumonia/) and (Wuhan or Hubei).tw,kw. |
| 7 | exp Influenza A Virus, H1N1 Subtype/ |
| 8 | (“A/H1N1*” or H1N1* or pdm09 or ((influenza or virus or pandemic) adj4 “2009”) or influenza A or swine flu).tw,kw. |
| 9 | 1 or 2 or 3 or 4 or 5 or 6 or 7 or 8 |
| 10 | ((respiratory adj3 (specimen* or sample* or swab*)) or sputum or nares or endotrachea* or endotrache* or endotra* or ((nasal or oral* or throat) adj3 (swab* or sample* or smear* or specimen*)) or NPS or OPS or ((endotrachea* or endotracheal*) adj2 aspirat*) or NPA or ETA or (deep adj4 saliva) or POS or “swab sample*” or “flocked swab*”).tw,kw. |
| 11 | Nasal cavity/ |
| 12 | Sputum/ |
| 13 | Nasopharynx/ |
| 14 | Oropharynx/ |
| 15 | *Saliva/ |
| 16 | Pharynx/ |
| 17 | (clinical adj2 (sample* or specimen*)).tw,kw. |
| 18 | (“RT-PCR” or “RTPCR” or “ddPCR” or “polymerase chain reaction”).tw,kw. |
| 19 | exp Coronavirus Infections/vi or exp Coronavirus/vi or exp Betacoronavirus/vi |
| 20 | polymerase chain reaction/ or multiplex polymerase chain reaction/ or real-time polymerase chain reaction/ or reverse transcriptase polymerase chain reaction/ |
| 21 | 10 or 11 or 12 or 13 or 14 or 15 or 16 or 17 or 18 or 19 or 20 |
| 22 | 9 and 21 |
| 23 | (vir* load* or vir* shed* or vir* burden or vir* titer* or vir* titre* or (vir* adj2 count*)).tw,kw. |
| 24 | (((copies or copy) adj2 (ml* or milli* or microl*)) or ((RNA* or vir*) adj2 concentration*)).tw,kw. |
| 25 | ((calibration adj1 curve*) or (standard adj1 curve*)).tw,kw. |

| | |
|---|---|
| 26 | ((ct* adj1 value*) or cycle threshold or (copies adj2 test*) or (copy adj2 test*) or ((copy or copies) adj2 number*)).tw,kw. |
| 27 | Viral load/ |
| 28 | Viral shedding/ |
| 29 | (“copy/m*” or “copies/m*” or “copy/test*” or “copies/test*”).tw,kw. |
| 30 | ((test or diagnos*) adj2 sensitiv*).tw,kw. |
| 31 | 23 or 24 or 25 or 26 or 27 or 28 or 29 or 30 |
| 32 | 22 and 31 |

**Supplement Table 4.** Search strategy used for Web of Science Core Collection.

| **Web of Science Core Collection** Nov 20, 2020 | |
|---|---|
| # | **Searches** |
| #1 | TOPIC: ((coronavirus* or "corona virus*" or betacoronavirus* or OC43 or NL63 or 229E or HKU1 or HcoV* or ncov* or covid* or "sars-cov*" or sarscov* or "Sars-coronavirus*" or "Severe Acute Respiratory Syndrome*" or "sudden acute respiratory syndrome*" or "2019-ncov*" or 2019nCov* or "2019-novel CoV" or corona or ((novel or new or nouveau) NEAR/2 (CoV or Pandemi*) ) OR (pneumonia and (Wuhan or Hubei) ) or "A/H1N1*" or H1N1* or pdm09 or ((influenza or virus or pandemic) NEAR/4 "2009") or "influenza A" or "swine flu"))<br>Indexes=SCI-EXPANDED, SSCI, A&HCI, CPCI-S, CPCI-SSH, ESCI Timespan=All years |
| #2 | TOPIC: (((respiratory NEAR/3 (specimen* or sample* or swab*) ) or sputum or nares or endotrachea* or endotrache* or endotra* or ((nasal or oral* or throat) NEAR/3 (swab* or sample* or smear* or specimen*) ) or NPS or OPS or ((endotrachea* or endotracheal*) NEAR/2 aspirat*) or NPA or ETA or (deep NEAR/4 saliva) or POS or "swab sample*" or "flocked swab*" or (clinical NEAR/2 (sample* or specimen*) ) or "RT-PCR" or "RTPCR" or "ddPCR" or "polymerase chain reaction"))<br>Indexes=SCI-EXPANDED, SSCI, A&HCI, CPCI-S, CPCI-SSH, ESCI Timespan=All years |
| #3 | TOPIC: (("vir* load*" or "vir* shed*" or "vir* burden" or "vir* titer*" or "vir* titre*" or (vir* NEAR/2 count*) or ((copies or copy) NEAR/2 (ml or mls or milli* or microl*) ) or ((RNA* or vir*) NEAR/2 concentration*) or (calibration NEAR/1 curve*) or (standard NEAR/1 curve*) or "ct value*" or "cycle threshold" or ((copies or copy) NEAR/2 test*) or ((copy or copies) NEAR/2 number*) or "copy/m*" or "copies/m*" or "copy/test*" or "copies/test*" or ((test or diagnos*) NEAR/2 sensitiv*)))<br>Indexes=SCI-EXPANDED, SSCI, A&HCI, CPCI-S, CPCI-SSH, ESCI Timespan=All years |
| #4 | #3 AND #2 AND #1<br>Refined by: [excluding] WEB OF SCIENCE CATEGORIES: ( VETERINARY SCIENCES )<br>Indexes=SCI-EXPANDED, SSCI, A&HCI, CPCI-S, CPCI-SSH, ESCI Timespan=All years |

**Supplement Table 5.** Search strategy used for medRxiv and bioRxiv.

| **medRxiv + bioRxiv (via Publish or Perish program)** Nov 20, 2020 |
| --- |
| **Keywords:** (covid OR coronavirus OR ncov OR hcov OR h1n1 OR “swine flu” OR COVID19 or SARS) AND (“copies/ml” OR “copy/ml” OR “viral load” OR “copies/test” OR “copy/test” OR “copies per ml” OR “copy per ml”)<br>**Publication name:** MedRxiv |
| **Keywords:** (covid OR coronavirus OR ncov OR hcov OR h1n1 OR “swine flu” OR COVID19 or SARS) AND (“copies/ml” OR “copy/ml” OR “viral load” OR “copies/test” OR “copy/test” OR “copies per ml” OR “copy per ml”)<br>**Publication name:** BioRxiv |

**Supplement Table 6.** Modified JBI critical appraisal checklist

Reviewer____________________________________Date____________________________

Author______________________________________Year________Record Number____________

| | Yes | No | Unclear | Not applicable |
|---|---|---|---|---|
| 1. Was the sample frame appropriate to address the target population? | □ | □ | □ | □ |
| 2. Were the study subjects and the setting described in detail? | □ | □ | □ | □ |
| 3. Did the study have consecutive inclusion of participants for case series and cohort studies? Did the study use probability-based sampling for cross-sectional studies? | □ | □ | □ | □ |
| 4. Was the response rate adequate, and if not, was the low response rate managed appropriately? | □ | □ | □ | □ |
| 5. Was the sample size adequate? | □ | □ | □ | □ |
| 6. Were valid methods used for the identification of the condition? | □ | □ | □ | □ |
| 7. Were standard, valid methods used for measurement of the exposure? | □ | □ | □ | □ |
| 8. Was the exposure measured in an objective, reliable way for all participants | □ | □ | □ | □ |
| 9. Was there clear reporting of clinical information of the participants? | □ | □ | □ | □ |
| 10. Was statistical analysis appropriate? | □ | □ | □ | □ |

Overall appraisal: Include □ Exclude □ Seek further info □

Comments (Including reason for exclusion)

______________________________________________________________________________

______________________________________________________________________________

______________________________________________________________________________

**Tool Guidance**

This modified checklist was based on a the JBI Critical Appraisal Checklists for case series, prevalence studies and analytical cross-sectional studies.

1. **Was the sample frame appropriate to address the target population?**

   This question relies upon knowledge of the broader characteristics of the population of interest and the geographical area.

   This study broadly investigates the respiratory viral load for the population of interest, which is the general population infected with SARS-CoV-2. The geographical area is not constrained. Sample frames restricted to particular subgroups within the general infected population were considered appropriate if they targeted one of the following groups analysed in our study: asymptomatic, presymptomatic, symptomatic, adult, pediatric, hospitalized, non-admitted, or community.

2. **Were the study subjects and the setting described in detail?**

   Certain diseases or conditions vary in prevalence across different geographic regions and populations (e.g. Women vs. Men, sociodemographic variables between countries). The study sample should be described in sufficient detail so that other researchers can determine if it is comparable to the population of interest to them

3. **Did the study have consecutive inclusion of participants for case series and cohort studies? Did the study use probability sampling for cross-sectional studies?**

   Inclusion of consecutive participants for case series and cohort studies yields results at lower risk of bias compared to other sampling methods for these study designs. Use of probability-based sampling methods for cross-sectional studies yields estimates at lower risk of bias compared to other sampling methods for this design. Studies that indicate a consecutive inclusion are more reliable than those that do not. For example, a case series that states ‘we included all patients (24) with osteosarcoma who presented to our clinic between March 2005 and June 2006’ is more reliable than a study that simply states ‘we report a case series of 24 people with osteosarcoma.’

4. **Was the response rate adequate, and if not, was the low response rate managed appropriately?**

   A large number of dropouts, refusals or “not founds” amongst selected subjects may diminish a study’s validity, as can a low response rates for survey studies. The authors should clearly discuss the response rate and any reasons for non-response and compare persons in the study to those not in the study, particularly with regards to their socio-demographic characteristics. If reasons for non-response appear to be unrelated to the outcome measured and the characteristics of non-responders are comparable to those who do respond in the study, the researchers may be able to justify a more modest response rate.

5. **Was the sample size adequate?**

The larger the sample, the narrower will be the confidence interval around the prevalence estimate, making the results more precise. An adequate sample size is important to ensure good precision of the final estimate. The sample size threshold was calculated as follows:

$$n^* = z^2\sigma / d^2$$

where $n^*$ is the sample size threshold, $z$ is the z-score for the level of confidence (95%), $\sigma$ is the standard deviation (assumed to be 3 $\log_{10}$ copies/ml, a fourth of the full range of rVLs) and $d$ is the marginal error (assumed to be 1 $\log_{10}$ copies/ml, based on the minimum detection limit for qRT-PCR across studies). This item was met if ≥75% of the included DFSO had ≥46 specimen measurements.

6. **Were valid methods used for the identification of the condition?**

Many health problems are not easily diagnosed or defined and some measures may not be capable of including or excluding appropriate levels or stages of the health problem. If the outcomes were assessed based on existing definitions or diagnostic criteria, then the answer to this question is likely to be yes. If the outcomes were assessed using observer reported, or self-reported scales, the risk of over- or under-reporting is increased, and objectivity is compromised. Importantly, determine if the measurement tools used were validated instruments as this has a significant impact on outcome assessment validity.

7. **Were standard, valid methods used for measurement of the exposure?**

The study should clearly describe the method of measurement of exposure. Assessing validity requires that a 'gold standard' is available to which the measure can be compared. The validity of exposure measurement usually relates to whether a current measure is appropriate or whether a measure of past exposure is needed.

In this study, standard methods to measure viral load in respiratory specimens are assays quantifying via one of the diagnostic sequences (*Ofr1b*, *N*, *RdRp* and *E* genes) for SARS-CoV-2.

8. **Was the exposure measured in an objective, reliable way for all participants?**

The study should clearly describe the procedural aspects of the measurement of exposure as well as factors that can contribute to heterogeneity in measurement.

In this study, objective, reliable interpretation of the exposure depends on the use of quantitative calibration; the specification of extraction; determination of the viral load as a standard metric (e.g., copies/ml or equivalent) or in a manner that can be converted to a standard metric; and, if present, specification of the amount of diluent (e.g., viral transport media) used.

9. **Was there clear reporting of clinical information of the participants?**

There should be clear reporting of clinical information of the participants such as the following information where relevant: disease status, comorbidities, stage of disease, previous interventions/treatment, results of diagnostic tests, etc.

In addition, there should be clear reporting of the number and types (asymptomatic, presymptomatic, symptomatic, adult, pediatric, hospitalized, non-admitted, community, etc.) of cases for measurements within the sampling periods of interest. For studies that include data outside of the infectious period, there should be clear reporting of clinical information for participants for the specimen measurements that were collected from within the infectious period.

10. **Was statistical analysis appropriate?**

As with any consideration of statistical analysis, consideration should be given to whether there was a more appropriate alternate statistical method that could have been used. The methods section of studies should be detailed enough for reviewers to identify which analytical techniques were used and whether these were suitable.

| **Risk of bias for each study** | |
|---|---|
| **Low** | The majority of critical appraisal criteria are met (≥6/10 items) and included item 1 (representative sample). The estimates are likely to be correct for the target population. |
| **High** | The majority of critical appraisal criteria are not met (<6/10 items) or did not include item 1 (representative sample). This may impact on the validity and reliability of the estimates. The estimates may not be correct for the target population. |
| **Unclear** | The majority of items are unclear. There was insufficient information to assess the risk of bias. |

### Respiratory viral loads during the infectious periods of SARS-CoV-2, SARS-CoV-1 and influenza A(H1N1)pdm09: a systematic review and meta-analysis

#### Citation

Paul Chen, Niklas Bobrovitz, Zahra Premji, David Fisman, Frank Gu. Respiratory viral loads during the infectious periods of SARS-CoV-2, SARS-CoV-1 and influenza A(H1N1)pdm09: a systematic review and meta-analysis. PROSPERO 2020 CRD42020204637 Available from:
https://www.crd.york.ac.uk/prospero/display_record.php?ID=CRD42020204637

#### Review question

What is the quantitative viral load in the respiratory tract of patients with SARS-CoV-2, SARS-CoV-1 or influenza A(H1N1)pdm09 during their respective infectious periods?

#### Searches

We will search without the use of filters or language restrictions the following sources: MEDLINE (via Ovid), EMBASE (via Ovid, Cochrane Central Register of Controlled Trials (CENTRAL) (via Ovid), Web of Science core collection ((including: Science Citation Index Expanded; Social Sciences Citation Index; Arts & Humanities Citation Index; Conference Proceedings Citation Index - Science; Conference Proceedings Citation Index - Social Sciences & Humanities; and Emerging Sources Citation Index), and MedRxiv & BioRxiv (both searched through Google Scholar via Publish or Perish). We will also search through the references of included articles.

A comprehensive search was developed by a librarian which included subject headings and keywords. The search strategy had 3 main concepts, disease, sample type, and outcome, and each concept was combined using the appropriate Boolean operators. The search was tested against a sample set of known articles that were pre-identified. The search results were exported from each database and uploaded to Covidence for deduplication and screening.

#### Types of study to be included

Include studies that (1) report viral loads of the viruses in human respiratory specimens; (2) report extractable data from the infectious periods of SARS-CoV-2 (-5 to +10 days from symptom onset for symptomatic patients, 0 to +10 days from diagnosis for asymptomatic patients), SARS-CoV-1 (during the symptomatic, or equivalent asymptomatic, period) and A(H1N1)pdm09 (-1 to +9 days from symptom onset for symptomatic patients, 0 to +9 days from diagnosis for asymptomatic patients); and (3) report data for two or more laboratory-confirmed cases.

Exclude studies that (1) report data for <2 people, including case reports, due to an increased risk of selection bias; (2) use only self-collected patient samples; (3) studies in animals or in vitro; (4) studies using environmental sampling; or (5) reviews of evidence (e.g., scoping, systematic, narrative).

We will include data from control groups in intervention studies, but we will not extract data for the intervention group.

#### Condition or domain being studied

COVID-19, SARS, and Influenza A(H1N1)pdm09

#### Participants/population

Inclusion: Humans of any age and symptomology diagnosed with COVID-19, SARS or Influenza A(H1N1)pdm09 based on RT-PCR for respiratory specimens in any setting.

Exclusion: Humans diagnosed using methods, including serological tests, not based on measuring viral load in respiratory specimens.

#### Intervention(s), exposure(s)

The exposure is COVID-19, SARS or Influenza A(H1N1)pdm09 confirmed by laboratory methods using respiratory specimens (i.e., nasopharyngeal swabs, oropharyngeal swabs, sputum, endotracheal aspirate, nasopharyngeal aspirate, posterior oropharyngeal saliva), irrespective of clinical signs and symptoms

#### Comparator(s)/control

The quantity of respiratory tract viral load will be compared between participants exposed to COVID-19, SARS and A(H1N1)pdm09.

#### Main outcome(s)

The main outcomes of this study are the mean and distribution of the respiratory viral load of SARS-CoV-2 throughout the infectious period compared with those of SARS-CoV-1 and influenza A(H1N1)pdm09.

##### * Measures of effect

Measures of effect:
(1) the mean and 95% confidence intervals for the respiratory viral load of each virus during the infectious period;

(2) the difference in means of the respiratory viral load of each virus during the infectious period;

(3) the distribution and descriptive statistics of respiratory viral loads for each virus during the infectious period;

(4) the difference in the distribution and descriptive statistics of respiratory viral loads for each virus during the infectious period.

#### Additional outcome(s)

The secondary outcomes of this study are the mean and distribution of the respiratory viral load of SARS-CoV-2 compared between each day (daily granularity) of the infectious period, compared between asymptomatic cases and symptomatic/presymptomatic cases and compared between adult and pediatric patients.

##### * Measures of effect

Measures of effect:

(1) the mean and 95% confidence intervals for the respiratory viral load of SARS-CoV-2 on each day of the infectious period;

(2) the difference in means of the respiratory viral load of SARS-CoV-2 between each day of the infectious period;

(3) the mean and 95% confidence intervals for the respiratory viral load of SARS-CoV-2 for asymptomatic cases and symptomatic/presymptomatic cases during the infectious period.

(4) the difference in means of the respiratory viral load of SARS-CoV-2 between asymptomatic cases and symptomatic/presymptomatic cases during the infectious period.

(5) the mean and 95% confidence intervals for the respiratory viral load of SARS-CoV-2 for adult patients and pediatric patients during the infectious period.

(6) the difference in means of the respiratory viral load of SARS-CoV-2 between adult patients and pediatric patients during the infectious period.

#### Data extraction (selection and coding)

Two reviewers will independently screened titles and abstracts, reviewed full texts, extract data and assess risk of bias. Screening will be conducted using the Covidence system. Numerical data will be extracted directly from the study into an excel file. Graphical data will be extracted using the Plot Digitizer software v4.3

(https://apps.automeris.io/wpd/). Inconsistencies will be resolved by consensus.

We will extract the following sets of data:

• Number of patients and specimens

• Characteristics of patients (e.g., age, sex, comorbidities, symptoms, setting of testing, geographical location)

• Type of respiratory specimen (i.e., nasopharyngeal swabs, oropharyngeal swabs, sputum, endotracheal aspirate, nasopharyngeal aspirate, posterior oropharyngeal saliva)

• Use and amount of dilutant (e.g., viral transport media)

• Quantitative viral load metrics (e.g., copies/ml, log10 copies/ml or an equivalent metric that can be converted to these metrics, such as Ct values when the appropriate calibration is included in the respective study)

Study investigators will be contacted to request missing data.

#### Risk of bias (quality) assessment

We will classify the design of included studies and assess their risk of bias using modified versions of critical appraisal tools developed by the Joanna Briggs Institute. Two reviewers will independently classify and assess each study using the tool. Disagreements will be resolved by consensus.

#### Strategy for data synthesis

To determine the respiratory viral loads of SARS-CoV-2, SARS-CoV-1 and A(H1N1)pdm09, data collected on positive viral loads in respiratory specimens will be converted to the viral concentration in the respiratory tract. For example, viral loads from swabbed specimens (nasopharyngeal swabs and oropharyngeal swabs) report the RNA concentration in viral transport media. Based on the uptake volume for specimen swabs (0.128 ± 0.031 ml, mean ± SD) and reported dilution in viral transport media in each study (e.g., 1 ml), we estimate the respiratory viral loads.

Means and 95% CI for the respiratory viral load during the entire infectious period from each study will be combined using random-effects meta-analysis. Heterogeneity will be reported using the $I^2$ statistic and p-value of the ?² test.

Individual patient data on each day of the infectious period will be combined to calculate the mean and 95% CI.

#### Analysis of subgroups or subsets

We will examine the main outcomes in the following sub-group analysis:

(1) age (adult >=18 vs. pediatric <18, or as defined in the studies)

(2) symptomatic/pauci-symptomatic/pre-symptomatic/ vs. asymptomatic patients

#### Contact details for further information

Paul Chen


#### Organisational affiliation of the review

University of Toronto
https://www.utoronto.ca/

#### Review team members and their organisational affiliations

Mr Paul Chen. University of Toronto
Dr Niklas Bobrovitz. Centre for Evidence-Based Medicine, University of Oxford, Oxford, United Kingdom;

Faculty of Medicine, University of Toronto, Toronto, Canada; Department of Critical Care Medicine, University of Calgary, Calgary, Canada
Dr Zahra Premji. Libraries and Cultural Resources, University of Calgary, Calgary, Canada
Professor David Fisman. Division of Epidemiology, Dalla Lana School of Public Health, University of Toronto, Toronto, Canada
Professor Frank Gu. Department of Chemical Engineering & Applied Chemistry, University of Toronto, Toronto, Canada

#### Type and method of review
Meta-analysis, Systematic review

#### Anticipated or actual start date
01 June 2020

#### Anticipated completion date
30 September 2020

#### Funding sources/sponsors
Natural Sciences and Engineering Research Council of Canada (NSERC); Toronto COVID-19 Action Fund

#### Conflicts of interest

#### Language
English

#### Country
Canada, England

#### Stage of review
Review Ongoing

#### Subject index terms status
Subject indexing assigned by CRD

#### Subject index terms
COVID-19; Communicable Diseases; Humans; Influenza A Virus, H1N1 Subtype; Influenza, Human; Severe Acute Respiratory Syndrome; Viral Load; severe acute respiratory syndrome coronavirus 2

#### Date of registration in PROSPERO
20 August 2020

#### Date of first submission
16 August 2020

#### Stage of review at time of this submission

| Stage | Started | Completed |
|---|---|---|
| Preliminary searches | Yes | Yes |
| Piloting of the study selection process | Yes | Yes |
| Formal screening of search results against eligibility criteria | Yes | No |
| Data extraction | No | No |
| Risk of bias (quality) assessment | No | No |
| Data analysis | No | No |

*The record owner confirms that the information they have supplied for this submission is accurate and complete and they understand that deliberate provision of inaccurate information or omission of data may be construed as scientific misconduct.*

*The record owner confirms that they will update the status of the review when it is completed and will add publication details in due course.*

#### Versions

20 August 2020

PROSPERO
This information has been provided by the named contact for this review. CRD has accepted this information in good faith and registered the review in PROSPERO. The registrant confirms that the information supplied for this submission is accurate and complete. CRD bears no responsibility or liability for the content of this registration record, any associated files or external websites.
